# Machine Learning Reveals Synovial Fibroblast Genes Associated with Pain Affect Sensory Nerve Growth in Rheumatoid Arthritis

**DOI:** 10.1101/2023.08.17.23294232

**Authors:** Zilong Bai, Nicholas Bartelo, Maryam Aslam, Caryn Hale, Nathalie E. Blachere, Salina Parveen, Edoardo Spolaore, Edward DiCarlo, Ellen Gravallese, Melanie H. Smith, Accelerating Medicines Partnership Program: Rheumatoid Arthritis and Systemic Lupus Erythematosus (AMP RA/SLE) Network, Mayu O. Frank, Caroline S. Jiang, Haotan Zhang, Myles J. Lewis, Shafaq Sikandar, Costantino Pitzalis, Anne-Marie Malfait, Rachel E. Miller, Fan Zhang, Susan Goodman, Robert Darnell, Fei Wang, Dana E. Orange

## Abstract

It has been presumed that rheumatoid arthritis (RA) joint pain is related to inflammation in the synovium; however, recent studies reveal that pain scores in patients do not correlate with synovial inflammation. We identified a module of 815 genes associated with pain, using a novel machine learning approach, Graph-based Gene expression Module Identification (GbGMI), in samples from patients with longstanding RA, but limited synovial inflammation at arthroplasty, and validated this finding in an independent cohort of synovial biopsy samples from early, untreated RA patients. Single-cell RNA-seq analyses indicated these genes were most robustly expressed by lining layer fibroblasts and receptor-ligand interaction analysis predicted robust lining layer fibroblast crosstalk with pain sensitive CGRP+ dorsal root ganglion sensory neurons. Netrin-4, which is abundantly expressed by lining fibroblasts and associated with pain, significantly increased the branching of pain-sensitive CGRP+ neurons *in vitro*. We conclude GbGMI is a useful method for identifying a module of genes that associate with a clinical feature of interest. Using this approach, we find that Netrin-4 is produced by synovial fibroblasts in the absence of inflammation and can enhance the outgrowth of CGRP+ pain sensitive nerve fibers.

**One Sentence Summary:** Machine Learning reveals synovial fibroblast genes related to pain affect sensory nerve growth in Rheumatoid Arthritis addresses unmet clinical need.

## INTRODUCTION

The four classic signs of inflammation are rubor, tumor, calor, and dolor – or redness, swelling, warmth, and pain. Inflammatory pain can be driven by cytokines, bradykinins, and prostanoids, which bind specific receptors on primary sensory neurons to cause heightened sensation of pain (*1*). Clinically, pain is not always proportional to inflammation and clinical scenarios in which pain is dissociated from inflammation are useful to study the non-inflammatory drivers of pain.

Rheumatoid arthritis (RA) is a chronic disease characterized by inflammation in the synovium, the tissue that lines the joint cavity. Despite great progress in developing an array of conventional synthetic, targeted synthetic, and biologic disease-modifying anti-rheumatic drugs (csDMARDs, ctDMARDs, and bDMARDs), which target relevant immune mediators (*2*), up to 20% of patients with RA are “difficult-to-treat”, that is they do not improve despite treatment with at least two bDMARD or tsDMARDs, with different mechanisms of action, after failing a csDMARD (*3*) It has been assumed that synovial inflammation is the cause of RA joint pain, however, recent studies have revealed that pain can be dissociated from inflammation in RA (*4–8*). Patients with RA and limited synovial inflammation, also known as “fibroid”, “low inflammatory” or “pauci-immune” synovium have as much pain as those with extreme inflammation (*9–11*). Patients with low inflammatory synovium tend to receive less benefit from treatment with anti-inflammatory drugs such as TNF inhibitors and disease-modifying anti-rheumatic drugs (DMARDs) (*12,13*), making their management particularly challenging for clinicians, and suggesting that traditional inflammatory pathways may not be the source of their discomfort.

Here, we performed a focused analysis on low inflammatory synovium to identify factors, beyond inflammation, that relate to and might mediate joint pain. Due to the small number of patients limiting the statistical power and patient reported outcome data being notoriously noisy, no existing machine learning approach is adequately powered to identify pain-associated gene modules from RA patients with low synovial inflammation. We developed a novel Machine Learning approach, called Graph-based Gene expression Module Identification (GbGMI). Using GbGMI, we discovered a group of 815 genes associated with pain in samples from patients with established RA but low synovial inflammation. We validated the pain-associated gene module on internal data and a second dataset of patients with early RA. Using sorted-cell subsets and single-cell RNA-seq data, we determined that lining fibroblasts express the majority of these pain-associated genes and, of these, we focused on genes predicted to interact with dorsal root ganglion sensory nerves. This led to the discovery that synovial lining fibroblasts produce Netrin-4 (Net4 or NTN4), which significantly augments branching of pain-sensitive CGRP+ sensory nerves *in vitro.* This work provides a novel approach to relate gene expression data to clinical data, uncovers a role for synovial fibroblast production of Netrin-4 in peripheral sensitization in RA, and nominates a panel of other targets that warrant additional study for their potential role in joint pain (Fig. S1).

## RESULTS

### Pain is not related to inflammation in RA patients with low inflammatory synovium

We categorized patients as high or low inflammatory using our previously reported histology scoring algorithm (*9*). Consistent with our prior studies, RA pain scores were not different between patients with high and low inflammatory synovium (Fig. 1A). Pain scores were associated with the level of synovial inflammation as measured by the density of cells per unit of tissue (cells/mm2) in patients with high inflammatory synovium, but not in patients with low inflammatory synovium (Fig. 1B).

**Fig. 1.**
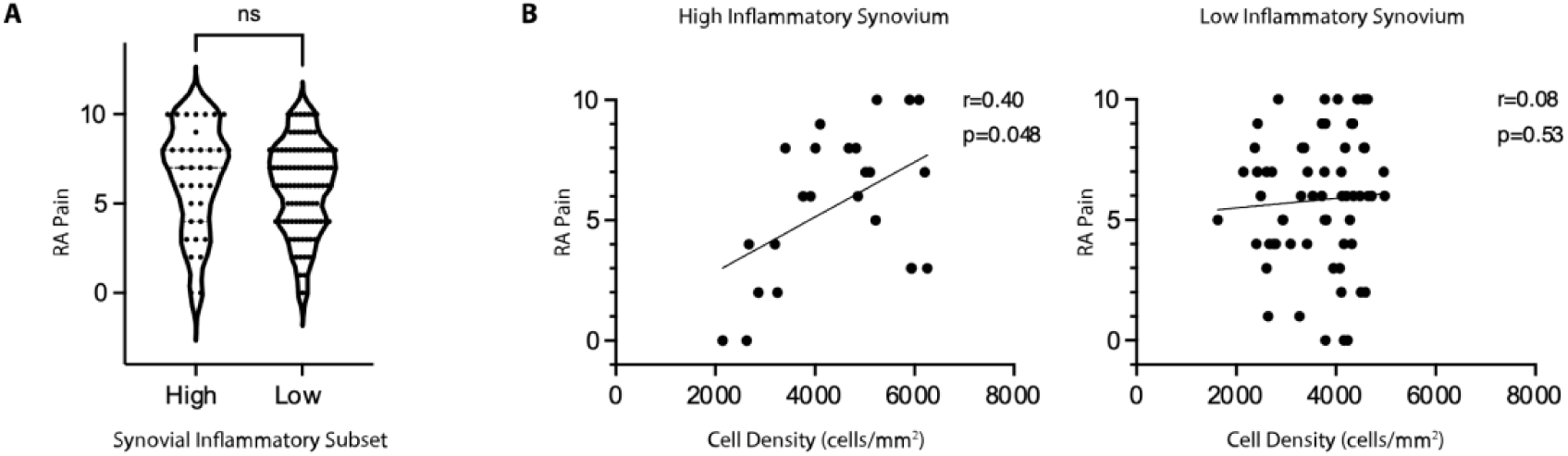
Pain is related to synovial inflammation in RA patients with high, but not low, synovial inflammation. (**A**) RA pain scores according to synovial tissue inflammatory classification in n=139 patients. (**B**) RA pain scores according to cell density, cells/mm^2^ of H&E (hematoxylin and eosin) stained synovial tissue, in samples classified as high or low inflammatory. ns=not significant in Mann Whitney test. r= Spearman’s rank correlation coefficient. p= two tailed p-value.

### GbGMI-identification of pain-associated synovial gene expression in patients with established RA

We first tested for genes that were significantly associated with pain using the usual RNA-seq analysis platform, limma (*14*). We failed to identify any significant individual genes that were correlated with pain suggesting the relationship of gene expression with pain could be multifactorial or nonlinear. We next hypothesized there might be groups of genes whose expression varies in association with pain. We developed an iterative machine-learning Graph-based Gene expression Module Identification (GbGMI) computational framework to uncover a group of genes whose expression is correlated with a given univariate clinical feature. Given a multi-modal input comprising a gene expression matrix *M* for *m* genes and *n* patients and an *n*-dimensional clinical feature vector *a*, GbGMI calculates the patient-to-patient similarity structure according to the given clinical feature, compares that to the gene expressions using the Laplacian score, and then determines the optimal number of genes that together associate with the clinical feature through statistical tests between the t-SNE-based summary scores of the selected genes and this clinical feature (see Materials and Methods. Fig. S2).

We first benchmarked this GbGMI approach by testing whether it would correctly identify genes known to be associated with inflammation as measured by cell density, which is associated with many significant individual genes as measured by limma (*14*). GbGMI identified a module of 2,713 genes whose gene expression summary score correlates with synovial tissue cell density (Fig. 2A-D). The positive control for this analysis was principal component one (PC1) of bulk synovial RNA-seq gene expression data, which was previously shown to associate with the extent of synovial inflammation and correlate significantly with synovial cell density (*15*), while the negative control was a gene expression summary score for a group of the top 5,000 most variably expressed genes. As expected, PC1 scores of gene expression were significantly correlated with synovial histologic cell density (Spearman ⍴=0.4, p=0.01) (Fig. 2F) while the gene expression summary score of the top 5000 most variably expressed genes were not (p=0.21) (Fig. 2E). The gene expression summary score of the GbGMI module of 2,713 genes had a further improved correlation to synovial histologic cell density (Spearman ⍴=0.59, p=0.0001) (Fig. 2G). Taken together, this analysis indicates that GbGMI is a useful method that outperformed PCA in identifying a module of genes that associate with the clinical feature of interest, synovial inflammation as measured by cell density.

**Fig. 2.**
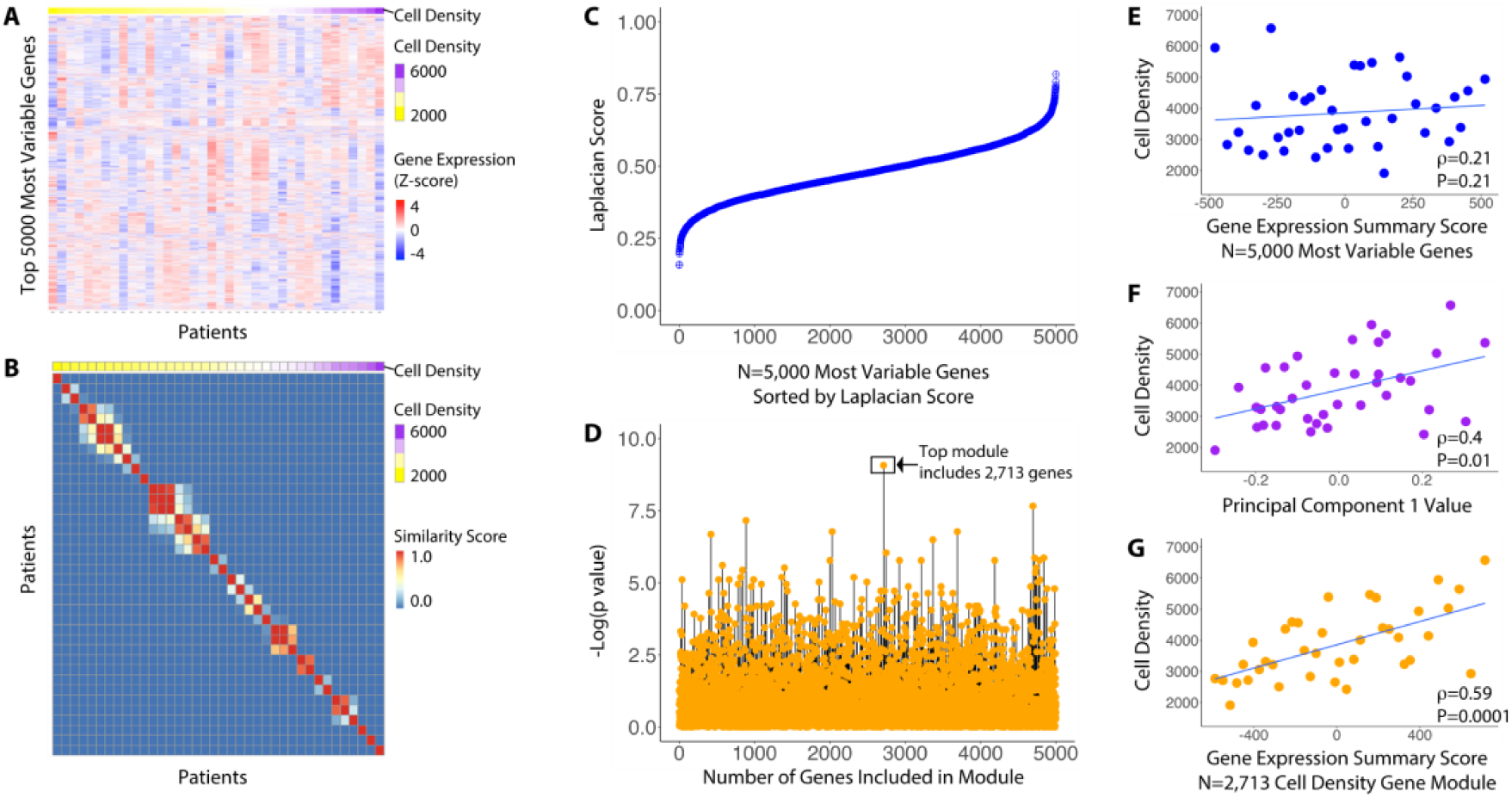
Identification of a synovial gene expression signature that correlates with synovial histologic cell density using GbGMI or PCA. (**A**) Expression heatmap of the top 5,000 most variably expressed genes in 38 patients. Gene expression levels (rows) are represented as z-scores for all patients. Patients (columns) are sorted by their mean nuclei densities. (**B**) Similarity matrix of synovial histologic cell densities. (**C**) Laplacian scores of the top 5,000 most variably expressed genes measuring how their expression levels varied compared to synovial histologic cell density similarity structure, each dot represents a gene, sorted by Laplacian score in ascending order. (**D**) -log(P value) of the correlation of the top-k-ranked groups of genes with nuclei density similarity structure, each dot represents a group of genes. (**E**) Synovial histologic cell density according to PC1 score of the top 5,000 most variably expressed genes in 38 patients. (**F**) Synovial histologic cell density according to the summary score of the 5,000 genes over the 38 patients. (**G**) Synovial histologic cell density according to the summary score of the 2,713 GbGMI-identified genes over the 38 patients. Statistics presented in **E**, **F**, and **G** indicate Spearman correlation coefficient and P-value.

We next applied GbGMI to define a module of genes associated with pain in patients with low inflammatory synovium. The majority of the 6,582 genes that distinguish high and low inflammatory synovium are increased in high inflammatory synovium and are enriched for pathways representing infiltrating immune cells. To uncover genes associated with pain, but not inflammation, we focused our analysis on 2,227 genes that were significantly increased in low inflammatory synovium relative to high inflammatory synovium (*9*) and on pain scores that document the extent of pain in the joint that was sampled (Hip Osteoarthritis Outcome Score/Knee Osteoarthritis Outcome Score (HOOS/KOOS) (Fig. 3A). The patient-reported pain scores *a* were transformed into a matrix of pairwise similarity scores between patients *S* (Fig. 3B). We next calculated the Laplacian score (*16*) for each of the 2,227 low-inflammatory genes based on its expression values (i.e., a row vector in *M*) and *S* (Fig. 3C). We then tested which number of top-ranked genes collectively best correlated with pain among RA patients with low synovial inflammation and identified an 815-gene module, which we refer to as the GbGMI-identified pain-associated genes (Fig. 3D). Although the summary score of all 2,227 low inflammatory genes did not correlate with pain, summary scores of the GbGMI-identified pain-associated genes were significantly correlated with the patient-reported HOOS/KOOS pain in patients with low inflammatory synovium (Fig. 3E). This correlation was not as pronounced when including all RA patients irrespective of inflammatory subset (Fig. 3F). Similar correlations were identified when the GbGMI-identified pain-associated genes were compared to Visual Analog Scores (VAS) report of pain (Fig. S3).

**Fig. 3.**
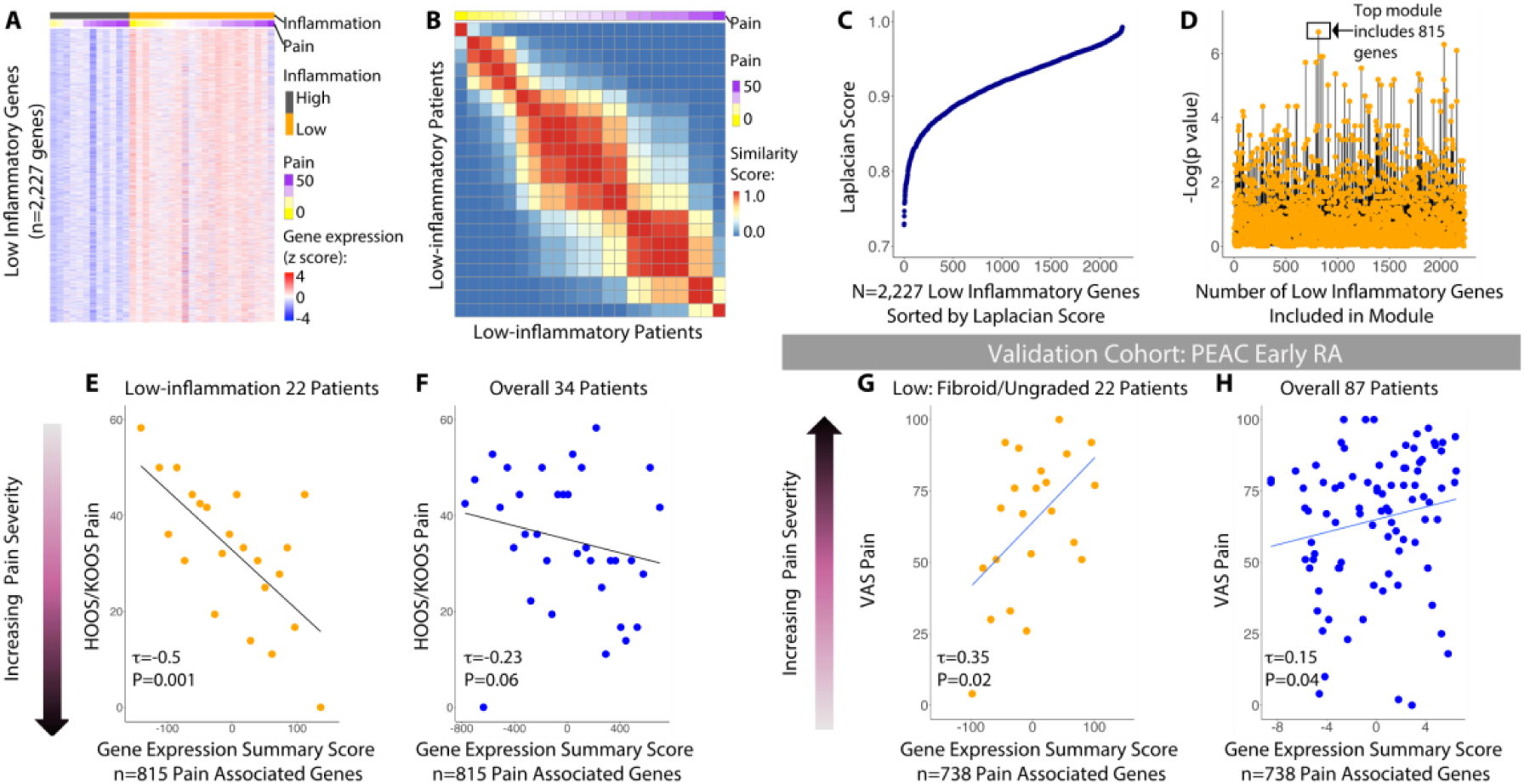
GbGMI-identification of pain-associated synovial gene expression in patients with established and early RA. (**A**) Expression heatmap of 2,227 genes with increased expression in low inflammatory synovium. Patients (columns) were grouped by inflammatory levels: high (n=12) versus low (n=22, low and mixed inflammatory subtypes identified by (*9*). (**B**) Similarity structure of HOOS/KOOS pain scores over patients. (**C**) Laplacian scores of the input 2,227 genes measure how their expression levels over patients relate to the pain-score-based similarity structure, each dot represents a gene, sorted by Laplacian score in ascending order. (**D**) Significance of the correlation between the top k ranked genes and HOOS/KOOS pain scores, each dot represents a group of genes. Established RA HOOS/KOOS pain score according to the summary score of the 815 GbGMI-identified genes in low inflammatory samples (**E**) or all samples, irrespective of inflammatory status (**F**). Early RA VAS pain scores, according to gene expression summary score of 738 GbGMI-identified pain-associated genes in patients with low inflammatory (fibroid or unassigned) synovial pathotype (**G**) or all patients, irrespective of synovial pathotype (**H**). Z-score is calculated from gene expression values over patients. Statistics presented in **E**, **F**, **G**, and **H** indicate Kendall correlation coefficient and P-value.

### GbGMI-identified pain-associated synovial gene expression in patients with early RA

Overfitting is a concern in using a graph-based machine-learning approach to identify groups of genes that associate with pain. It is possible that the GbGMI-identified pain-associated genes correlate with pain in the dataset in which they were discovered, but not in other external datasets. We next sought to test whether the pain-associated gene module identified in patients with established disease was also associated with pain in a second, independent Pathobiology of Early Arthritis Cohort (PEAC) dataset (*10*) of synovial biopsy samples from patients with early (mean of 6 months of symptoms), untreated, RA. 2,018 of the 2,227 low inflammatory genes and 738 of the 815 pain-associated genes discovered in the established RA were also measured in this dataset. The 738 GbGMI-identified pain-associated genes were also significantly correlated with Visual Analog Score (VAS) pain in patients with early RA with low (fibroid or undefined) inflammatory synovium (Fig. 3G), while the 2,018 low-inflammatory genes were not. In this early RA cohort, the 738 GbGMI-identified pain-associated genes were also associated with pain in all patients, irrespective of synovial inflammatory subset, as well, though the association was again not as robust as was seen in those with low inflammatory synovium (Fig. 3H). Of note, the range of GbGMI summary scores decreased when all samples were included. The association of the GbGMI-identified genes with pain was robust in the low inflammatory samples, but persisted even when all patients were included, suggesting that these genes may play a role, albeit less pronounced, in pain in high inflammatory synovium, where inflammatory mediators are highly likely to also contribute.

### GbGMI-identified pain-associated genes are enriched with neurogenesis pathways and predominantly expressed by synovial fibroblasts

We next sought to understand the biological meaning and the direction of the association of the 815 GbGMI genes with pain in RA patients with low synovial inflammation. Limma was performed to detect genes whose expression correlates with pain and genes were ranked by limma according to this correlation. Of note, though limma did not identify any significant (FDR <0.05) individual genes correlated with pain (Fig. S4A), as a group, expression of the 815 GbGMI-identified pain genes was significantly decreased as the HOOS/KOOS pain score increased (adjusted p-value: 7.38e-12 (ks.test) (Fig. S4B and S4C), indicating a positive correlation with pain severity. The 815 pain-associated genes included ephrin (EPHA3, EPHA6, EPHA7) and semaphorin (SEMA3B, SEMA3E, SEMA4C, SEMA5A, SEMA6D) family members and were significantly enriched in nervous system development and neuron projection pathways. On the other hand, the 1,412 non-pain associated genes included CD55, PRG4, CSPG4, MERTK, genes known to be involved in the normal functions of lining macrophages and fibroblasts (*17*), and were enriched in molecular function and rRNA processing, but not neuron projection pathways (Fig. 4A). We next examined which cells express the GbGMI pain-associated genes by comparing their expression in sorted bulk synovial B cells, fibroblasts, monocytes, and T cells, which offers high depth coverage of RNA but less cell type resolution, and sorted single cells, which offers higher cell type resolution to cell subtypes but less depth of coverage, from the Accelerating Medicines Partnership dataset (*18*). Comparison of the pain-associated genes across sorted bulk synovial B cells, fibroblasts, monocytes, and T cells indicated fibroblasts express the highest levels of pain-associated genes (Fig. 4B and 4C). We reasoned pain associated genes might be more robustly expressed in fibroblasts because of a relative enrichment in fibroblasts, compared to immune cells, in low-inflammatory samples.

**Fig. 4.**
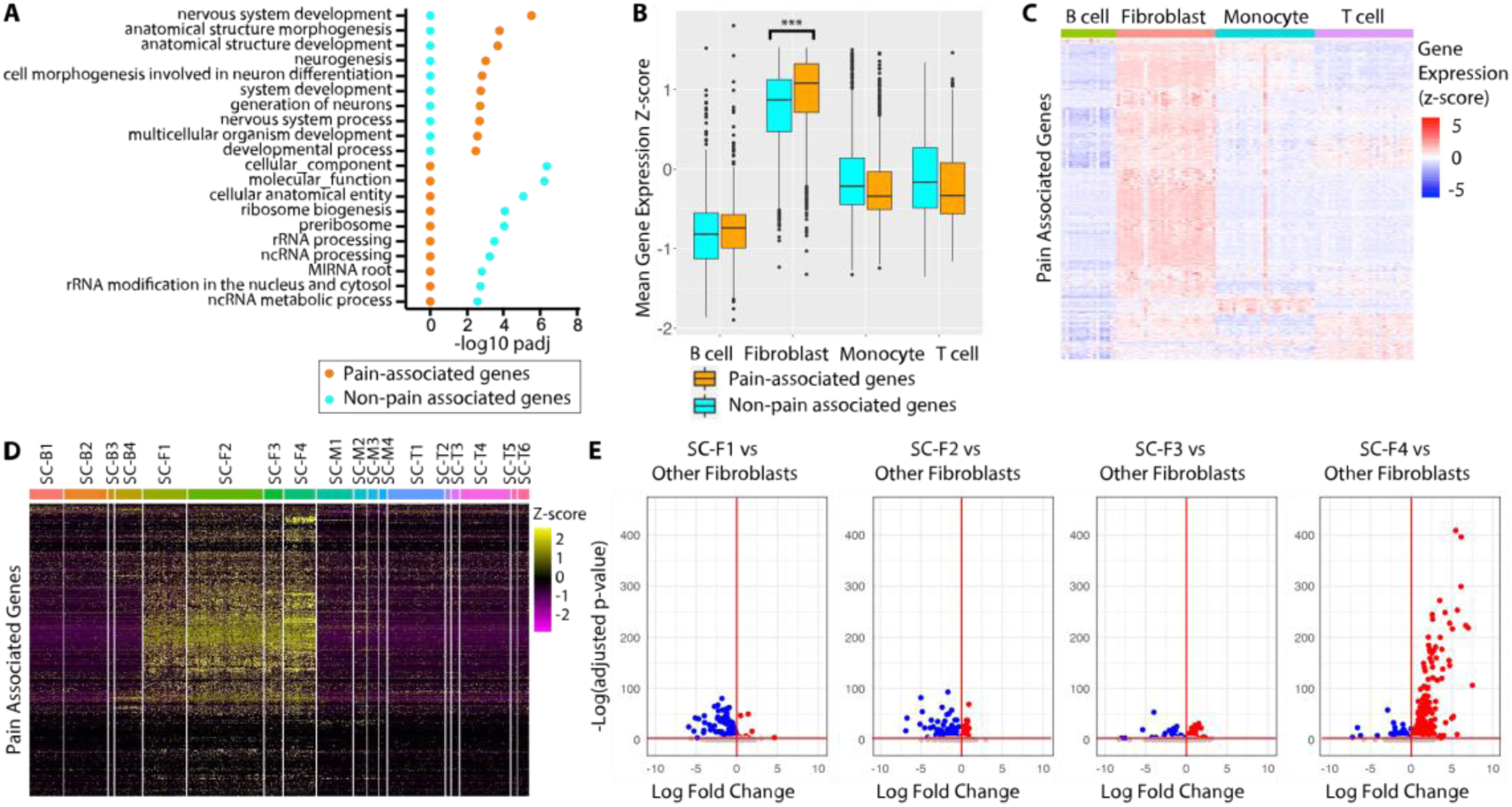
GbGMI-identified pain-associated gene signature is expressed by synovial lining layer fibroblasts. (**A**) g:Profiler pathway enrichment analysis of 815 pain associated genes and 1,412 non-pain associated low inflammatory genes. (**B**) Mean gene expression z score of pain-associated genes and non-pain-associated genes detected in sorted bulk B cells (CD45+CD3−CD19+), fibroblasts (CD45−CD31−PDPN+), monocytes (CD45+CD14+), and T cells (CD45+CD3+). Z score is calculated on the basis of TPM normalized counts. *** indicates p-value < 0.001. (**C**) Per sample gene expression z score of 769 pain-associated genes detected in sorted cell types from **B**. (**D**) Expression heatmap of 797 pain genes with non-zero variance in expression values across a subset (n=4,354) of RA synovial cells in 18 unique cell populations (of B cells: SC-B1-4, Fibroblasts: SC-F1-4, Monocytes: SC-M1-4, T cells: SC-T1-6), which were identified from the 5,265 scRNA-seq profiles by an integrated analysis based on CCA from the Accelerating Medicine Partnership (*18*). Z score is calculated on the basis of log2(CPM+1) transformed UMIs counts over the RA synovial cells. (**E**) Volcano plots of 794 pain genes in scRNA-seq profiles (Immport Accession #SDY998) (*11*) with non-zero variance in expression values across the subset (n=1,532) of RA synovial fibroblasts in three sublining subsets, CD34+ (SC-F1), HLA-DRAhi (SC-F2), and DKK3+ (SC-F3) and one lining subset (SC-F4). Each volcano plot shows the differential expression analysis (using Seurat function FindMarkers) of the genes in each RA synovial fibroblast subtype compared to the other three, where x-axis shows *log*_2_(Fold Change) and y-axis - log(adjusted p-value). The significantly increased, significantly decreased, and non-significantly differentially expressed genes are indicated by different colors. The horizontal and vertical red lines respectively indicate the threshold of significance [-log(adjusted p-value = 0.05)] and the separation threshold between increased and decreased gene expression *log*_2_(Fold Change) = 0.

However, when looking only at fibroblasts, the pain-associated genes were significantly increased compared to the non-pain associated genes (Fig. 4B). The marked differences in pathways enriched in pain-associated and non-pain associated genes as well as the difference in relative expression levels within fibroblasts indicates the GbGMI method did not select a random group of fibroblast genes. Further analysis of the single cell RNA-seq dataset also confirmed that the fibroblast subsets express the highest levels of pain-associated genes (Fig. 4D). Gene expression analysis among fibroblast subsets indicated that, compared to the other fibroblast subsets, lining CD55+ fibroblasts (SC-F4) express the highest level of GbGMI-identified pain-associated genes (Fig. 4E). Taken together, these analyses indicate GbGMI-identified pain associated genes are expressed by synovial fibroblasts and enriched for neurogenesis pathways.

### Predicted interactions between lining fibroblasts and dorsal root ganglion neurons

Given the pain-associated genes were enriched in neuron projection pathways, we next explored predicted interactions of pain-associated synovial fibroblast genes with dorsal root ganglion (DRG) sensory neurons, which contain the soma of synovial innervating nociceptive neurons.

We performed receptor-ligand interaction analysis to identify predicted receptor-ligand pairs using the pain-associated genes expressed by four synovial fibroblast subtypes in human RA synovial tissue and genes expressed in a human DRG bulk RNA-seq dataset (*19, 20*). Lining fibroblasts (SC-F4) were predicted to have the highest number of ligand-receptor interactions (57 SC-F4 ligands to hDRG receptors) (Fig. 5A and 5B). To further clarify which types of DRG nerves might be predicted to interact with synovial fibroblasts, we also performed receptor-ligand interaction analysis between the pain-associated genes expressed by the synovial fibroblast subsets (*18*) and a mouse scRNA-seq DRG dataset (*21*). Again, lining fibroblasts (SC-F4) were predicted to have the highest number of receptor-ligand interactions and were predicted to interact with proprioceptors, as well as A-*b*, A-*d* and CGRP peptidergic neurons (Fig. 5C and table S3). Comparison of the expression of 21 ligand or receptor encoding pain-associated genes of SC-F4 revealed a gradient of pain-associated genes that are relatively lowly expressed in SC-F1 cells and most highly expressed in SC-F4 cells, with HBEGF, CTGF, and NTN4 among the most robustly expressed (Fig. 5D).

**Fig. 5.**
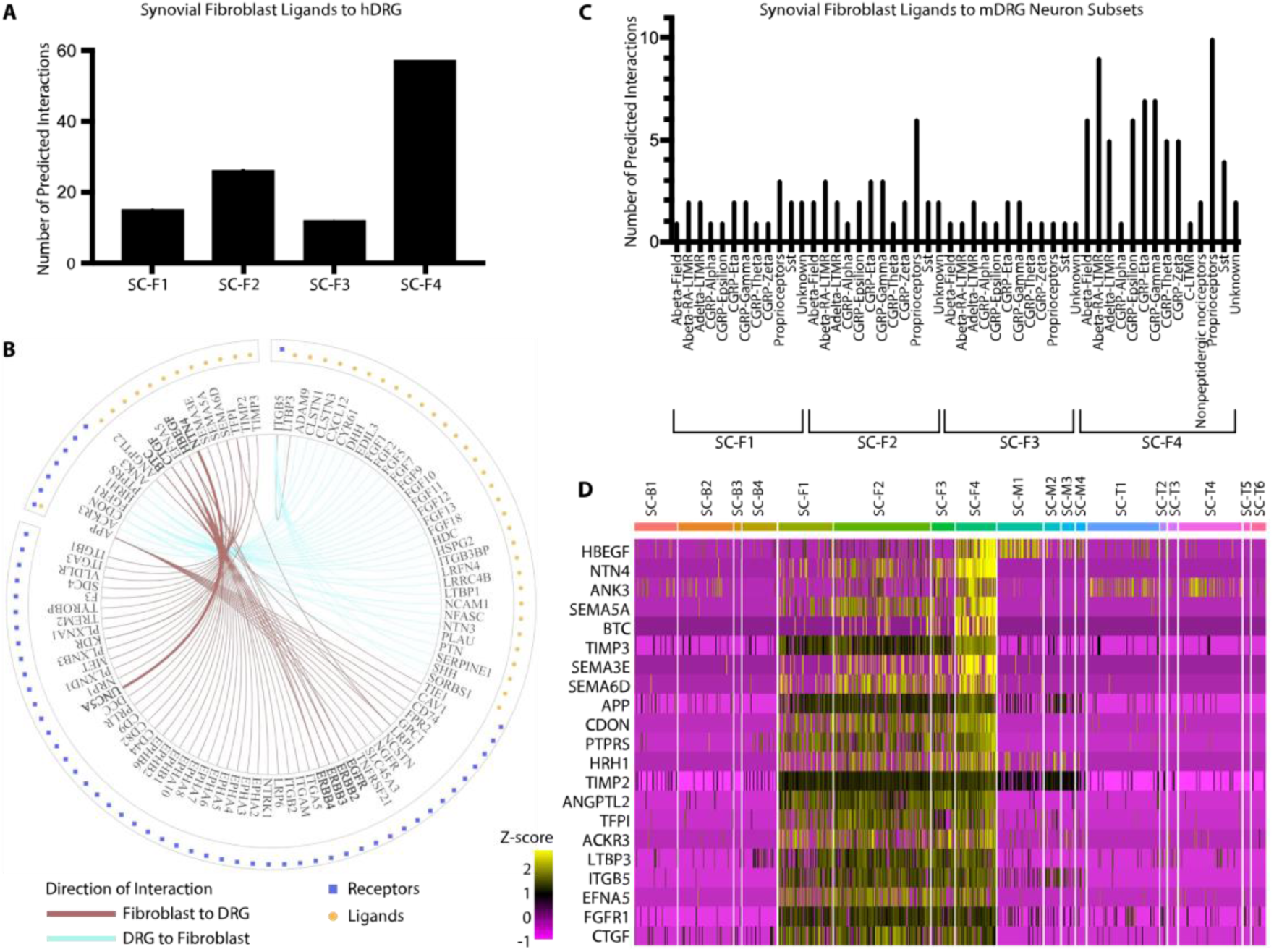
Filtering on synovial fibroblast genes predicted to influence dorsal root ganglion sensory nerves. (**A**) Number of predicted synovial fibroblast ligands with paired DRG receptors according to fibroblast subset. (**B**) Predicted ligand-receptor pairs between synovial lining fibroblasts (SC-F4) and DRG tissue. The outermost circle indicates the source of expression, synovial lining fibroblast SC-F4 or DRG tissue. The dots represent whether the gene is ligand-coding or receptor-coding. The inner layer contains gene names. The color of the lines connecting the gene names indicate the direction of the interaction. (**C**) Number of predicted interactions between synovial fibroblast subsets and various DRG subsets. (**D**) Expression heatmap of 21 pain-associated ligand/receptor encoding marker genes of synovial lining fibroblast cells with non-zero variance in expression values across a subset (n=4,354) of RA synovial cells in 18 unique cell populations (of B cells: SC-B1-4, Fibroblasts: SC-F1-4, Monocytes: SC-M1-4, T cells: SC-T1-6), which were identified from the 5,265 scRNA-seq profiles by an integrated analysis based on CCA from the Accelerating Medicine Partnership (*18*). Z-score is calculated on the basis of log2(CPM+1) transformed UMIs counts over the RA synovial cells. The genes are ranked top down by their log fold change in DEA of lining fibroblast vs other fibroblasts.

### Products of synovial fibroblasts influence adult dorsal root ganglion sprouting and branching in response to injury

We next sought to test whether any of the pain-associated synovial fibroblast genes, discovered in this analysis, might directly influence the growth of pain sensitive neurons in the synovium. Unmyelinated, small, CGRP+ nerve fibers are responsible for pain transmission and can be found in the synovium extending up to the lining layer (*22*), indicating synovial lining fibroblast interaction with CGRP+ nociceptive fibers could take place within relatively close range. While pathway analysis indicated many of the pain associated genes are associated with neurogenesis, the majority of this data was defined in studies of the central nervous system of developing embyros of model organisms, not adult human diseased joint tissue. *NTN4* was of interest because it was associated with pain in this dataset and is highly expressed by synovial fibroblasts. Netrin-4 (NTN4 or Net4) is a secreted member of the netrin family of proteins, of which, Netrin-1, is the first discovered and best described for its role as an axon attractant during embryogenesis (*23*). Though Net4 has only 30% sequence homology to Netrin-1, it has been shown to augment embryonic olfactory bulb sprouting and thalamocortical branching (*24–26*). It is not known whether or how Netrin-4 might influence injured adult DRG CGRP+ pain sensitive neurons. We cultured adult mouse dissociated DRG neurons with either no supplements, nerve growth factor (Ngf), as a positive control, since it is known to produce robust effects on axon sprouting (*27,28*), or Net4, and measured survival, sprouting, and branching of CGRP+ DRG neurons. CGRP status of the DRG neurons was assigned using immunoflourescent stains (Fig. S4). While there was no effect of either Ngf or Net4 on CGRP+ neuron survival (Fig. 6A), or sprouting (Fig. 6B), compared to untreated CGRP+ neurons, Net4 significantly augmented branching *in vitro* (Fig. 6B).

**Fig. 6.**
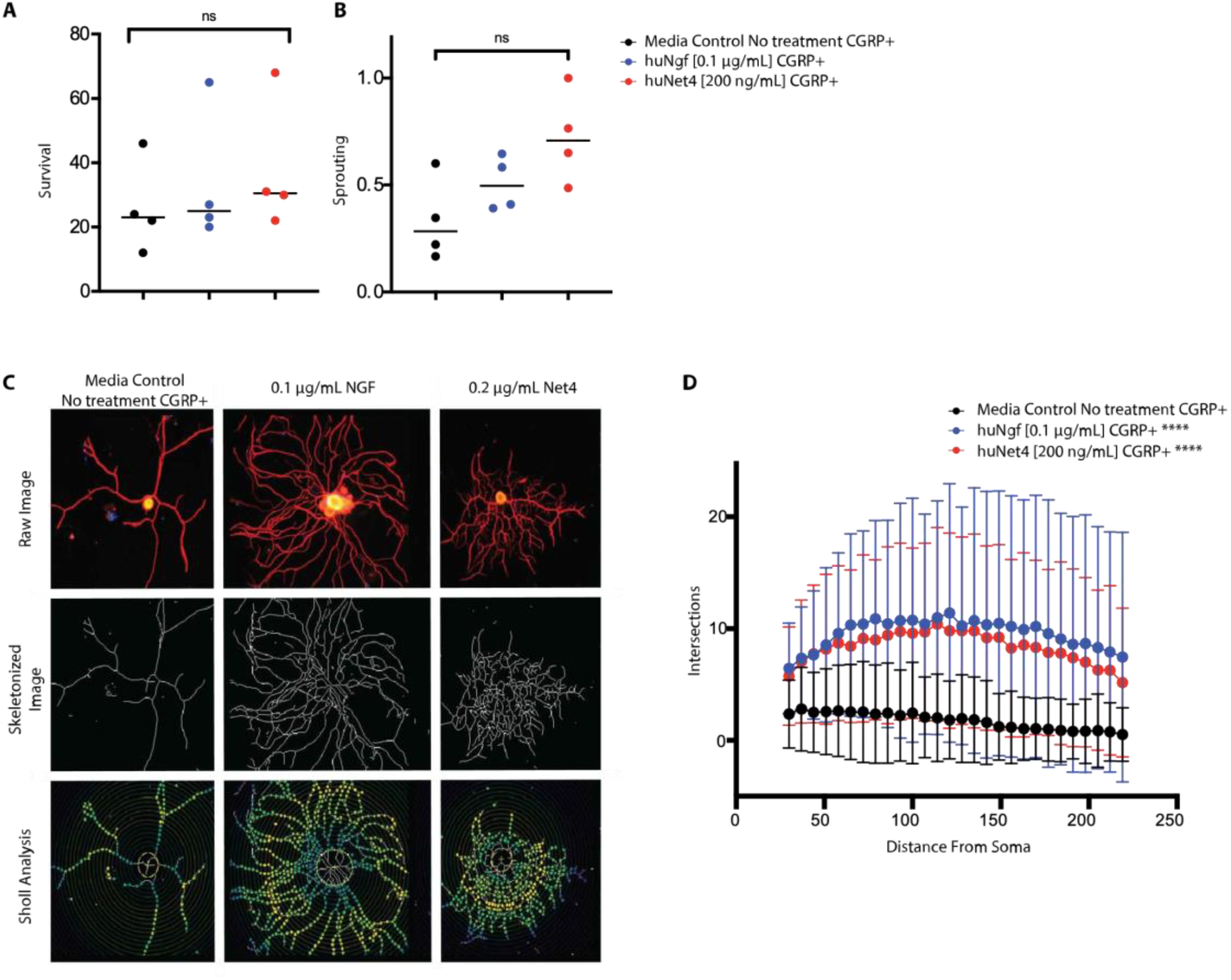
Effects of synovial fibroblasts products on dorsal root ganglion neurons in vitro. (**A**) Survival of CGRP+ DRG neurons cultured with media alone, Ngf, or Net4. Survival was measured by the number of Map2b+B3tub+ cells >10μm. Each dot represents the sum of five 10x magnification views from one experiment. Data from 4 experiments are presented. ns indicates not significant in Kruskal-Wallis test. (**B**) Sum of sprouting neurons divided by the total number of neurons cultured with media alone, Ngf, or Net4. Neurons with at least three axon branches greater than two times the size of the soma were classified as sprouting. Each dot represents the sum of five 10x magnification views from one experiment. Data from 4 experiments are presented. ns indicates not significant in Kruskal-Wallis test. (**C**) Representative images of Sholl analysis of branching of DRG neurons cultured with media alone (no treatment), Ngf, or Net4. (**D**) Branching, as measured by the number of shell intersections of neurites, in DRG neurons cultured with media alone (no treatment), Ngf, or Net4. Each dot represents the median with confidence interval of 40 neurons imaged from four experiments (10 neurons per experiment). **** indicates p<0.0001 in two way ANOVA group*radius interaction with post-hoc Dunnett’s multiple comparisons of each treatment group to the no treatment group.

## DISCUSSION

Using our newly developed GbGMI approach, we discovered a module of 815 genes, whose expression correlates with patient report of pain and are enriched in neuron projection pathways. These pain-associated genes are most robustly expressed by lining layer fibroblasts. The proteins encoded by these pain-associated genes include netrins, semaphorins, and ephrins, proteins that ensure neurons and their axons develop in the correct stages and places required for normal nervous system development. Human knee innervating sensory neurons extend their pseudounipolar axons approximately one meter from their origin in the DRG to their destination in the joint. A complex interplay of multiple molecules at distinct concentrations attract or repel neurons to tune their projections in space and time during development (*29*). Our findings that RA synovial fibroblasts express an array of neural guidance genes that associate with patient report of pain and augment CGRP+ nociceptor growth *in vitro* are consistent with recent studies in osteoarthritis (OA), which identified aberrant CGRP+ sensory neurite sprouting into normally aneural cartilage (*30–33*). While RA tends to affect all three knee compartments similarly, OA tends to affect the medial side more severely. Compared to synovial fibroblasts from the nonpainful side, synovial fibroblast conditioned media from the painful side of knee OA increased neuron survival and longest branch length *in vitro* (*34*). Our studies suggest there may be a mechanism, in both RA and OA, by which synovial fibroblasts can play a role in mediating joint pain. Our studies also led to the discovery that Netrin-4, which has been shown to affect thalamocortical axon sprouting in embryogenesis(*26*), augments branching of injured pain-sensitive CGRP+ nerves *in vitro*. Netrin-4 binds the extracellular matrix molecule, laminin, specifically laminin ***γ***1, encoded by the gene LAMC1, and in doing so, dramatically weakens matrix stiffness (*25*). Laminins are large glycoproteins that are abundant in synovium and LAMA4 and LAMC1 expression are increased in RA synovium. It is possible that a compliant extracellular matrix facilitates CGRP+ neurite sprouting or branching. Netrin-4 also binds cell surface receptors, such as Unc5B (*26*), which are expressed on several dorsal root ganglion. Avenues for future research include *in vivo* animal studies to test the function of NTN4, as well as other identified pain-associated genes, and explore their potential in drug or treatment design.

It is worth noting that there are many other genes associated with patient report of pain in this dataset that warrant additional study such as HBEGF, BTC, and CTGF, which augment neuronal sprouting in response to injury and have been used to treat pathological pain alleviation in other clinical scenarios (*35,36*). In addition, neural guidance molecules also have important functions outside the nervous system, such as affecting angiogenesis, lung branching morphogenesis, immunomodulation, and tumorogenesis, and these non-neuronal effects of this family of molecules warrant further study in the context of arthritis.

In summary, here we present GbGMI, a feature-selection method that can be used to identify pathology-specific multi-gene signatures by exploiting patient-to-patient similarity structures in different feature spaces when the number of patients is limited. We used GbGMI to identify pain-associated genes expressed by synovial fibroblasts in low-inflammatory RA synovial tissues, leading to the discovery that synovial fibroblast-sensory neuron crosstalk may relate non-inflammatory joint pain and providing an avenue to pursue novel targets for the management of joint pain.

## MATERIALS AND METHODS

### Study Approval

This study includes data from 102 RA patients undergoing arthroplasty at the Hospital for Special Surgery (HSS) in New York. All patients met either the American College of Rheumatology (ACR)/European League Against Rheumatism 2010 Classification criteria (*37*) and/or the ACR 1987 criteria for RA. Patient data including Knee disability and Osteoarthritis Outcome Score (KOOS) (*38*) and Hip disability and Osteoarthritis Outcome Score (HOOS) (*39*) questionnaire were collected. RA pain scores indicate response to the question “How much pain have you felt due to your rheumatoid arthritis during the last week?”, with responses ranging from 1 to 10. Condition pain scores indicate response to the question, “How much pain have you had because of your condition OVER THE PAST WEEK? Please indicate how severe your pain has been” with responses ranging from 1 to 10. This study was approved by the HSS Institutional Review Board (approval no. 2014-233), the Rockefeller University Institutional Review Board (approval no. DOR0822), and the Biomedical Research Alliance of New York (approval no. 15-08-114-385). All participating patients provided their signed informed consent.

### H&E Histologic Scoring

Synovial samples were obtained from the most grossly inflamed (dull and opaque) area of synovium. If there were no obviously inflamed areas, samples were obtained from standard locations: the femoral aspects of the medial and lateral gutters and the central supratrochlear region of the suprapatellar pouch. Each tissue biopsy was sectioned at 5-micrometer thickness and stained with Harris modified hematoxylin solution and eosin Y (H&E) manufactured by Epredia in Kalamazoo, MI. An expert musculoskeletal pathologist scored fourteen synovial histologic features in a single section for each patient: lymphocytic inflammation, mucoid change, fibrosis, fibrin, germinal centers, lining hyperplasia, neutrophils, detritus, plasma cells, binucleated plasma cells, Russell bodies, sub-lining giant cells, synovial lining giant cells, and mast cells. Detailed methods for scoring these features are described in prior studies (*9,40*) and available at www.hss.edu/pathology-synovitis, the classification algorithm is available at Immport www.immport-open/public/study/displayStudyDetail/SDY1299.

### Gene Expression Established Arthritis Cohort

RNA was extracted from 39 bulk synovial tissue samples collected from an established RA cohort and previously sequenced as described in (*41*) (ImmPort Accession #SDY1299). In brief, these libraries were prepared using TruSeq messenger RNA (mRNA) Stranded Library kits, 50-bp paired-end reads were sequenced on a HiSeq2500 platform, and reads were aligned to hg19 using STAR (*42*). Samples with >0.1% globin mRNA were excluded from further analysis. After quality control, ComBaT in the Bioconductor SVA package (*43*) was used for batch effect correction, and DESeq2 (*44*) was used to normalize the data. Consensus clustering identified three gene expression clusters characterized by different levels of synovial inflammation: low (n=14), intermediate (n=11), and high (n=14) (*9*). Of these 39 RA patients, 38 had mean nuclei density data for the benchmarking analysis (*15*), and 26 had low inflammatory synovium, for which all 26 had scores for lymphocytic inflammation and lining hyperplasia and 22 had scores of fibrosis and HOOS/KOOS pain scores. We used limma (*14*) to test for gene expression correlates of fibrosis, lymphocytic inflammation, lining hyperplasia and pain.

### Gene-set enrichment analysis (GSEA)

A moderated t-test was performed between each gene expression profile and the pain scores over the subset (n=22) low-inflammatory patients. The resulting log2 fold-change therefore offers a score of correlation between the expression of each gene and the pain. The moderated t-statistic (ratio of log2-fold change to its standard error) was used to rank the genes. The fgsea R package (*45*) was used for enrichment analysis of the GbGMI-identified subset (n=815) of genes associated with pain as well as the set of marker genes for each of the 18 synovial cell subpopulations (200 marker genes per subpopulation) (*11,18*) (ImmPort Accession #SDY998). Since the HOOS/KOOS pain scores are from 0 (worst joint health) to 100 (best joint health) (*38,39,46*), a gene set more correlated to the bottom of the list hence negatively correlated with HOOS/KOOS pain scores, indicating positive correlation with pain.

### Gene Expression Early Arthritis Cohort

For external validation of the pain-associated genes identified by our method, we downloaded the bulk RNA-seq of early RA patients from the Pathobiology of Early Arthritis Cohort (PEAC) on ArrayExpress (https://www.ebi.ac.uk/arrayexpress/experiments/E-MTAB-6141/samples/)

Using only sample with sufficient data quality, reads were pseudoaligned to hg38 with Kallisto in order to generate a counts matrix. Counts were normalized for read depth (counts into log2(CPM+1) (Counts Per Million)) and batch corrected using removeBatchEffect from limma.

### Gene Expression in bulk sorted cells and single cells

In the analyses related to RA synovial cell types, we used the sorted-population bulk RNA-seq gene expression data of the synovial T cells (CD45+, CD3+, CD14–), monocytes (CD45+, CD3–, CD14+), B cells (CD45+, CD3–, CD14–, CD19+), and fibroblasts (CD45–, CD31–, PDPN+) collected by fluorescence activated cell sorting (BD FACSAria Fusion) directly in buffer RLT (Qiagen) (*11,18*). Normalized read counts per gene as transcripts per million (TPMs) were used (ImmPort Accession #SDY998) (*11*).

We used the single-cell RNA-seq data preprocessed by (*18*), where the gene expression levels were quantified by counting UMIs (Unique Molecular Identifiers) and transformed into log2(CPM + 1). From the scRNA-seq profiles for 32,391 genes and 5,265 cells left after rigorous quality control, 18 unique cell populations were identified by an integrated strategy based on canonical correlation analysis. Specifically, in fibroblasts: CD34+ sublining fibroblasts (SC-F1), HLA-DRAhi sublining fibroblasts (SC-F2), DKK3+ sublining fibroblasts (SC-F3), and CD55+ lining fibroblasts (SC-F4); in monocytes: IL1B+ pro-inflammatory monocytes (SC-M1), NUPR1+ monocytes (SC-M2), C1QA+ monocytes (SC-M3), and interferon (IFN) activated monocytes (SC-M4); in T cells: three CD4+ clusters: CCR7+ T cells (SC-T1), FOXP3+ regulatory T cells (Treg cells) (SC-T2), and PDCD1+ TPH and TFH (SC-T3) cells, and three CD8+ clusters, GZMK+ T cells (SC-T4), GNLY+GZMB+ cytotoxic lymphocytes (CTLs) (SC-T5), and GZMK+GZMB+ T cells (SC-T6); in B cells: naïve IGHD+CD27– (SC-B1), IGHG3+CD27+ memory B cells (SC-B2), autoimmune-associated B cell (ABC) cluster (SC-B3) with high expression of ITGAX (also known as CD11c) and a plasmablast cluster (SC-B4) with high expression of immunoglobulin genes and XBP1 (*18*). The top 200 marker genes for each subpopulation identified by differential expression analysis in the original publication (*18*) were used as signature genes for individual cell-types within RA synovium.

### Description of our Graph-based Gene expression Module Identification (GbGMI) framework

We developed GbGMI, a graph-based machine learning framework of algorithms, to identify a gene expression module that strongly correlates with pain in low-inflammatory RA synovial tissues (Fig. S2). We use *M* ∈ *R*^*m*×*n*^ to represent the input gene expression matrix of genes and patients. We use a numeric vector *a* ∈ *R*^*n*^ to represent the pain scores reported by these patients. We quantified the quality *Q* of a selected gene subset via the correlation between their collective expression and the pain score, and then searched for a gene subset with optimal quality *Q* as a feature selection task (*47–50*). As exhaustive search through all possible subsets of an input gene set to optimize *Q* is computationally intractable (*51*), we adapted and integrated the feature scoring strategy used in the filter feature-selection/ranking approaches (*52–54*) and the feature subset scoring strategy used in the wrapper feature-selection approaches (*50,55*). Specifically, we generated a gene prioritization list ℒ_*s*_ according to how well each individual gene expression respects the geometric structure over patients built according to their pain scores. Then, the quality *Q*_*k*_ of the *k*-th candidate gene subset comprising the top *k* genes in ℒ_*s*_ is evaluated, for *k* ranging from 1 to *m*. The first *k*^∗^ where *Q*_*k*_ peaked is used as the cut-off point on ℒ_*s*_. This subset of *k*^∗^ genes is the output pain-associated gene module.

#### Gene prioritization with adapted Laplacian Score algorithm

(*16*) Given the input gene expression matrix and pain-score vector, we ranked the genes by the way each gene expression vector (i.e., a row vector in the gene expression matrix *M*) respected a given geometric structure over the patients encoded in an *n* × *n* similarity matrix *S* based on the pain score vector *a* instead of assuming independent observations in the correlation tests (Fig. S2, step A). To compute *S*, a Gaussian kernel, which empirically outperformed other types of kernels (*56,57*), was adopted to map the Euclidean distance between the pain scores of each pair of patients and into a similarity measure *S*(*i*, *j*) ∈ [0,1]:

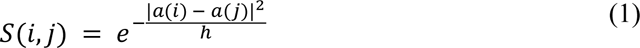

where *h* corresponds to the bandwidth or smoothing factor in a kernel metric definition. This formulation forces the similarity measure between any two patients with significantly different pain scores to be close to 0, while pushing the similarity measure between the patients of pain scores within a certain range (depending on the smoothing factor) to be closer to 1. This promotes the *locality* we want to focus on. We set *h* to control the local neighborhood of patients on graph *S* according to the theoretical range of the pain scores (e.g., *h* = 100 for HSS HOOS/KOOS pain score or PEAC VAS characteristic since either ranges between 0 and 100).

The Laplacian Score of each gene is then computed to evaluate how well this gene’s expression on these patients preserves *S*. (Fig. S2, step B.) This is different from the original publication (*16,58*) which aimed to preserve the input feature space (e.g., the input low-inflammatory genes), wherein we aim to select features (e.g., the *k*^∗^ pain-associated genes). The Laplacian matrix of *S* is defined by *L* = *D* − *S*, where *D* is a diagonal matrix with *D*_*ii*_ indicating the degree of node (i.e., patient) *i* in the weighted graph *S* (i.e., *D* = *diag*(*S* 1)). For the *r*-th gene, let *M*_*r*,∗_ be its *n*-dimensional gene expression vector across the patients (i.e., the *r*-th row vector in the gene expression matrix *M*), its Laplacian Score *ls*(*r*) is computed as:

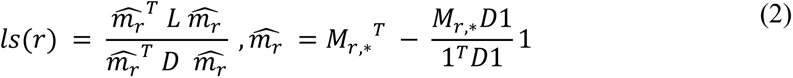

where the symbol 1 denotes a column vector whose all elements are 1’s with dimensionality determined by context. The gene prioritization list ℒ_*s*_ is generated by sorting the input *m* genes according to their Laplacian Scores in ascending order. The smaller the Laplacian Score of a gene, the better its expression data respect the geometric structure defined by *S* over the patients (according to objective function analysis (*16*)). Each top-*k* subset on ℒ_*s*_ forms a candidate gene subset for selection. The rows of *M* were reordered according to ℒ_*s*_. (Fig. S2, step C.)

#### Quantification of candidate ***k***-gene subset quality ***Q***_***k***_

*The **Q***_***k***_ *of the **k**-th candidate* gene subset is measured based on the association between their collective expression pattern and the pain-score vector *a*. Given the gene expression submatrix *M*_1:*k*,∗_ of this candidate *k*-gene subset, ***Q***_***k***_ is computed through two steps: Firstly, project the *k*-gene expression vector of each patient into a univariate summary score that preserves the patient-to-patient similarity structure in the original *k*-dimensional feature (gene) space. This addresses the dimensionality mismatch between the multi-gene expression and the univariate patient-level pain score (*9*). The resulting summary score vector of the *n* patients is denoted ***s***_***k***_. (Fig. S2, step D.) Secondly, quantify ***Q***_***k***_ by using the statistical significance of correlation test (e.g., the −*log*(*p*-value) of Kendall’s correlation test) between *s*_*k*_ and *a* over the same patients. We chose the first *k*^∗^, where ***Q***_***k***_∗ peaked, as the cut-off point on the sorted gene prioritization list ℒ_*s*_. This subset of *k*^∗^ genes is the pain-associated gene module identified by our GbGMI framework. (Fig. S2, step E.)

For Fig. S2, step D, We used the t-distributed Stochastic Neighborhood Embedding (t-SNE) (*57*). The resulting summary score vector *s*_*k*_ for the *n* patients respect how the gene expression data was arranged in the *k*-gene feature space of this candidate gene subset. We hereon briefly sketch the instantiation of t-SNE with the variables involved in our study. Following the SNE framework (*59*), the directional similarity of patient *j* to patient *i* based on their multi-gene expression vectors *M*_1:*k*,*i*_ and *M*_1:*k*,*j*_ is :

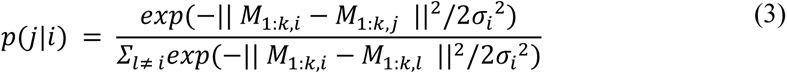

where the variance of the Gaussian kernel *σ*_*i*_^2^ is chosen such that the perplexity of the conditional probability distribution over all points *j* ≠ *i* defined by

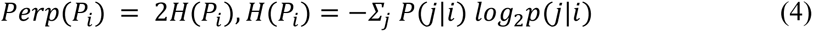

matches a pre-specified value. The perplexity in this context can be interpreted as an estimation about the number of close neighbors of each patient on graph *S*. Therefore, we specify the perplexity based on the rounded mean degree of the similarity graph *S* built from *a*. The symmetric SNE was used for mathematical and computational convenience in the t-SNE formulation by defining the following undirected similarities:

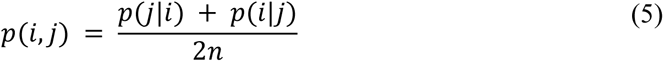

where *n* is the number of patients. Since *Σ*_*i*,*j*_*p*(*i*, *j*) = 1, this is a valid probability distribution on the set of all pairs (i,j). The t-SNE step in this algorithm uses the t-distribution with one degree of freedom (also known as Cauchy distribution) as the one-dimensional similarity kernel applied to pairs of summary scores defined by:

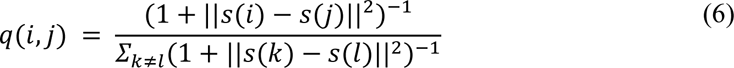

The main idea of this t-SNE-based step is to arrange the patients in a one-dimensional space such that the similarities *q*(*i*, *j*) between *s*(*i*) and *s*(*j*) match *p*(*i*, *j*) as close as possible in terms of the Kullback-Leibler (KL) divergence. Thus the loss function is:

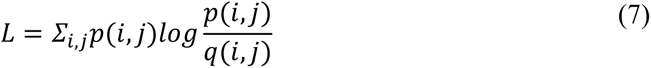

A summary score vector *s*_*k*_ is computed for each candidate *k*-gene subset, rendering a set of *m* such vectors {*s*_*k*_| *k* = 1,2, ⋯, *m*}.

Our framework GbGMI can be applied to identify a subset of genes that collectively associate with some other patient-level numeric attribute beyond the main focus of this paper (the HOOS/KOOS pain score). Our framework is also flexible and adaptive in different contexts by replacing specific computational components with other design choices (e.g., other embeddings instead of t-SNE for computing summary scores of selected gene expression).

### Pathway Enrichment Analysis

We used g:Profiler (https://biit.cs.ut.ee/gprofiler/gost) (*60*) for enrichment analysis of genes in the KEGG (*61*), REACTOME (*62*), WikiPathways pathways (*63*), and the Gene Ontology (GO): molecular functions (MF), cellular components (CC) and biological processes (BP) gene sets (*64*). A hypergeometric test was performed to estimate statistical significance, and all P values were adjusted for multiple testing using g:SCS (Set Counts and Sizes) correction (*65*). The genes from the bulk RNA-seq data (ImmPort Accession #1299) (*9*) were used to custom the statistical domain scope (i.e., background gene list), the size of which describes the total number of genes used for random selection and is one of the four parameters for the hypergeometric probability function for computing statistical significance used in g:GOSt (*60*).

### Ligand-receptor Interaction Analysis

For predicting potential ligand-receptor interactions between RA synovium and DRG, we used the normalized counts per gene as transcripts per million (TPMs) reported in a prior study where human DRG tissue samples were sequenced using bulk RNA-seq and analyzed (*20*). We used the scRNA-seq data from a mouse DRG dataset (*21*) to generate the transcriptome profiles of individual cell-types within DRG. Our analysis used the expression values and metadata for each subpopulation of component cells provided in the National Center for Biotechnology Information (NCBI) Gene Expression Omnibus (GEO) databases (#GSE139088) generated by the original publication. The cell subpopulations in this mDRG scRNA-seq dataset are specifically 15 somatosensory neuron subtypes (Aβ field/SA1, Aβ RA-LTMR, Aδ-LTMR, C-LTMR, CGRP-α, CGRP-ε, CGRP-η, CGRP-γ, CGRP-θ, CGRP-ζ, Nonpeptidergic nociceptors, TrpM8, Proprioceptors, SST, Cold thermoceptors) and a cluster of unassigned/non-neuronal cells (*21*).

For ligand-receptor analysis, we used the ligand-receptor pair interactome containing more than 3,000 interactions (denoted *I*^*o*^) created by a previous publication (*19*). This prior model mapped the potential ligand-receptor interactions between DRG neurons and distinct cell types within tissues throughout the human body by curating a database of ligand and receptor pairs across the genome, based on the literature and curated bioinformatics databases (*66–71*).

To identify the interactome for a specific ordered pair of tissue or cell types, the ligand-encoding genes from *I*^*o*^ were intersected with the genes that satisfy the ligand-side inclusion criteria in the tissue or cell type wherein we investigate the upstream component of the signaling. Similarly the receptor-encoding genes of *I*^*o*^ were intersected with the genes satisfying the receptor-side inclusion criteria in the tissue or cell type wherein we investigate the downstream component of the signaling. We generated the interactomes in two different contexts: (i) between each of the 4 fibroblast subtypes in human RA synovial tissue and the human DRG (hDRG) (*20*) and (ii) between each of the 4 synovial fibroblast subtypes and each of the 14 neuronal cell types of mouse DRG (mDRG) (*21, 41*) (GEO GSE139088). We set the filtering criteria to consider a confined subset of genes from the bulk or single-cell RNA-seq data in a specific tissue or cell type depending on the data available and the question being asked in each context. In particular:

For each fibroblast subtype in human RA synovial tissue, we included its top 200 marker genes (*18*) intersected with the GbGMI-identified 815 pain-associated genes. From the hDRG bulk RNA-seq data, we included the genes consistently expressed (> 0.1 TPM) in the 16 human DRG samples with “yes” for their associated pain status (20). On the mDRG scRNA-seq data, we included the differentially expressed genes (adjusted p-value< 0.05) identified by the FindMarkers function from the R package Seurat (*72*) for each of the 15 neuronal cell subpopulations compared to the rest. Lastly, the ligand(/receptor)-coding genes with 0 corresponding receptor(/ligand)-coding genes were excluded from the built interactome.

We also performed ligand-receptor interaction analysis between the pain-associated genes expressed by the four synovial fibroblast subtypes and a scRNA-seq dataset of adolescent mouse nervous system (MNS) (*73*). We predicted considerable connections between lining fibroblasts (SC-F4) and peripheral sensory neurofilament, peptidergic and non-peptidergic neurons, as well as sympathetic cholinergic and noradrenergic neurons (Fig. S6). These analyses are detailed in Supplementary Materials.

### DRG Dissection and Digestion

Prior to extraction of the DRG neurons, chambered coverslips (ibidi 80286) were coated overnight at 37°C with poly-l-lysine (Sigma-Aldrich P4832) and then for two hours at 37°C with mouse sarcoma basement membrane laminin (Sigma-Aldrich L2020) diluted 1:50 with 1XPBS and, with one also coated with 0.2μg/mL hNet4 (R&D Systems 1254-N4).

For each assay, sensory DRG neurons were harvested from two female 6-8 week old C57BL/6 mice (Jackson Laboratories), under a dissection microscope using forceps and placed into a waiting 15mL conical tube on ice containing 1X L-15 medium (ThermoFisher 21083027). DRGs were spun down at 950 rpm for two minutes. Media was aspirated and replaced with 1mL L-15 containing 10mg/mL Dispase II (Sigma-Aldrich 04942078001) and 10mg/mL Collagenase IV (ThermoFisher 17104019). DRGs were then placed at 37°C for 20 minutes. Enzyme solution was then carefully aspirated and replaced with 2mL L-15. Pellet was resuspended thoroughly with a 1000ul pipette. 25ul 10mg/mL DNAse I was then added and once again the cells were placed at 37°C for 20 minutes. The cells were then spun down for 5 minutes at 950 rpm and resuspended in 5mL L-15. After another 5 minute centrifugation, the cells were resuspended in 1mL L-15 and layered on top of 15% ice-cold BSA (Sigma-Aldrich A7906) and spun down at room temperature for 8 minutes at 1179 rpm to remove myelin. The cell pellet was resuspended in 1X Neurobasal Plus media (ThermoFisher A3582901) containing B27 (ThermoFisher 17504001) diluted 1:50, glutaMAX (ThermoFisher 35050061) diluted 1:100, and gentamicin sulfate (Abbott Laboratories), and then plated onto the pre-coated slides. 0.1ug/mL human beta-NGF (R&D Systems 256-GF) was added to the media of the positive control slide chamber.

### Immunofluorescence Staining

After 24 hours, cells were fixed with 4% PFA (Electron Microscopy Sciences 15714-S) for 10 minutes at room temperature and then permeabilized with 0.2% Triton on ice. They were then blocked with 3% BSA for 1 hour at RT, then incubated at 4°C overnight with primary antibodies rabbit anti-CGRP (Immunostar 24112) diluted 1:20,000 and mouse anti-beta III tubulin (abcam ab78078) diluted 1:1000 in 3% BSA. The next day, cells were incubated with secondary antibodies goat anti-Mouse Alexa Fluor 555 (ThermoFisher A21422) and goat anti-Rabbit Alexa Fluor 488 (ThermoFisher A11008), both diluted 1:1000, for two hours at room temperature.

Following the second incubation, plates were washed three times in 1XPBS, with the second wash containing DAPI (ThermoFisher D1306) diluted 1:1000.

### Neurite Imaging, Quantification, and Comparison

Each chambered slide was examined under a Keyence fluorescence microscope and the accompanying software was used to capture a stitched image of a large 5x5 area randomly selected on the plate at 10X magnification. Five of these images were captured per plate. To quantify sprouting versus non-sprouting DRG neurons, each image was examined in ImageJ and used the Cell Counter plugin to keep track of the total number of sprouting neurons, identified as having at least three neurites extended from the cell body where the extensions were more than twice the diameter of the cell body and exhibited some degree of branching. From across these images, n=10 neurons that exhibited branching were selected and their branching was quantified using the Sholl Analysis plugin. Each analysis used a start radius of 30 pixels, an end radius of 225 pixels, and a step size of 7. Mixed model repeated measures analysis was used to analyze Sholl data. The model included group, radius (categorical), and group*radius interaction as fixed effects. A significant group*radius interaction indicated group differences and branching.

## Data Availability

All data produced in the present work are contained in the manuscript.

https://www.immport.org/shared/study/SDY1299

https://www.immport.org/shared/study/SDY998

## List of Supplementary Materials

### Materials and Methods

Identify differentially expressed genes via ANOVA test on pain-level-based groupings

Ablation study of GbGMI using alternative approaches for gene prioritization

Sensitivity analysis of GbGMI on the HSS low-inflammatory patients

Methodological Validation of GbGMI on the early RA dataset

Ligand-receptor analysis using mouse nervous system scRNA-seq

**Fig S1** Overview of Study.

**Fig S2** A diagram illustrating the framework of GbGMI for identifying a synovial gene expression signature that associates with pain.

**Fig S3** GbGMI-identified pain-associated synovial gene expression compared against VAS pain scores in patients with established RA at different levels of synovial inflammation.

**Fig S4** GbGMI-identified pain-associated gene expression increases with increasing pain severity.

**Fig S5** Representative staining of CGRP+ DRG neuron *in vitro*.

**Fig S6** Sensitivity analysis of GbGMI on the HSS low-inflammatory patients.

**Fig S7** Potential ligand-receptor interactions between synovial fibroblasts and neurons in the mouse nervous system.

**Tables S1** Pathway Analysis

**Tables S2** Fibroblast–DRG receptor ligand interaction analysis

**Tables S3** Fibroblast-Sensory nerve subset receptor ligand interaction analysis

**Data file S1** Data file S1 Pathway Analysis.xlsx

**Data file S2** Data file S2 Fibroblast - DRG receptor ligand interaction analysis.xlsx

**Data file S3** Data file S3 Fibroblast-Sensory nerve subset receptor ligand interaction analysis.xlsx

## Acknowledgments

We thank the participants who provided synovial tissue and blood samples for this study. We thank **Alison North, Ph.D. and Christina Pyrgaki, Ph.D.** from the Rockefeller University’s Bio-Imaging Resource Center, RRID:SCR_017791, for help with imaging and image analysis. This project was funded by the Arthritis Foundation, the Block Family Foundation as well as NIH R01 AR078268 (DEO), UC2AR081025 (DEO, SG), UL1 TR001866 (DEO), R01AR077019 (REM), R01AR064251 (AM), R01AR060364 (AM), P30AR079206 (AM) and the Accelerating Medicines Partnership Program: Rheumatoid Arthritis and Systemic Lupus Erythematosus (AMP RA/SLE) Network. The AMP Program is a public-private partnership that includes AbbVie Inc., the Arthritis Foundation, Bristol-Myers Squibb Company, the Foundation for the National Institutes of Health, GlaxoSmithKline, Janssen Research and Development, LLC, the Lupus Foundation of America, the Lupus Research Alliance, Merck Sharp & Dohme Corp., the National Institute of Allergy and Infectious Diseases, the National Institute of Arthritis and Musculoskeletal and Skin Diseases, Pfizer Inc., the Rheumatology Research Foundation, Sanofi, and Takeda Pharmaceuticals International, Inc. Funding for AMP RA/SLE work was provided through grants from the National Institutes of Health (UH2-AR067676, UH2-AR067677, UH2-AR067679, UH2-AR067681, UH2-AR067685, UH2-AR067688, UH2-AR067689, UH2-AR067690, UH2-AR067691, UH2-AR067694, and UM2-AR067678). The work of ZB and FW was supported by NSF 1750326. The PEAC study was supported by funding from the UK Medical Research Council (MRC) [grant number G0800648] and core work was supported by grants from Versus Arthritis [Experimental Arthritis Treatment Centre, grant number 20022].

## Funding

Arthritis Foundation

Block Family Foundation

National Institutes of Health grant R01 AR078268 (DEO)

National Institutes of Health grant UC2AR081025 (DEO, SG)

National Institutes of Health grant UL1 TR001866 (DEO)

National Institutes of Health grant R01AR077019 (REM)

National Institutes of Health grant R01AR064251 (AM)

National Institutes of Health grant R01AR060364 (AM)

National Institutes of Health grant P30AR079206 (AM)

Accelerating Medicines Partnership Program: Rheumatoid Arthritis and Systemic Lupus Erythematosus (AMP RA/SLE)

Network

National Institutes of Health grant UH2-AR067676 (AMP RA/SLE work)

National Institutes of Health grant UH2-AR067677 (AMP RA/SLE work)

National Institutes of Health grant UH2-AR067679 (AMP RA/SLE work)

National Institutes of Health grant UH2-AR067681 (AMP RA/SLE work)

National Institutes of Health grant UH2-AR067685 (AMP RA/SLE work)

National Institutes of Health grant UH2-AR067688 (AMP RA/SLE work)

National Institutes of Health grant UH2-AR067689 (AMP RA/SLE work)

National Institutes of Health grant UH2-AR067690 (AMP RA/SLE work)

National Institutes of Health grant UH2-AR067691 (AMP RA/SLE work)

National Institutes of Health grant UH2-AR067694 (AMP RA/SLE work)

National Institutes of Health grant UM2-AR067678 (AMP RA/SLE work)

National Science Foundation grant 1750326 (ZB, FW)

UK Medical Research Council (MRC) grant G0800648 (PEAC study)

Versus Arthritis [Experimental Arthritis Treatment Centre, grant 20022] (PEAC study core work)

**Accelerating Medicines Partnership Program: Rheumatoid Arthritis and Systemic Lupus Erythematosus (AMP RA/SLE) Network includes:**

Jennifer Albrecht^8^, Jennifer H. Anolik^8^, William Apruzzese^9^, Brendan F. Boyce^8^, David L. Boyle^10^, Michael B. Brenner^9^, S. Louis Bridges Jr.^3^, Christopher D. Buckley^11^, Jane H. Buckner^12^, Vivian P. Bykerk^1,3^, James Dolan^9^, Laura T. Donlin^1,3^, Thomas M. Eisenhaure^13^, Andrew Filer^11^, Gary S. Firestein^10^, Chamith Y. Fonseka^9,13^, Ellen M. Gravallese^14^, Peter K. Gregersen^15^, Joel M. Guthridge^16^, Maria Gutierrez-Arcelus^9,13^, Nir Hacohen^13^, V. Michael Holers^7^, Laura B. Hughes^17^, Lionel B. Ivashkiv^1,3,18^, Eddie A. James^12^, Judith A. James ^16^, A. Helena Jonsson^9^, Stephen Kelly^19^, James A. Lederer^9^, Yvonne C. Lee^20^, David J. Lieb^13^, Arthur M. Mandelin II^20^, Mandy J. McGeachy^21^, Michael A. McNamara^1,3^, Joseph R. Mears^9,13^, Nida Meednu^8^, Larry Moreland^21^, Harris Perlman^20^, Javier Rangel-Moreno^8^, Deepak A. Rao^9^, Soumya Raychaudhuri^9,13,23^, Christopher Ritchlin^8^, William H. Robinson^24^, Mina Rohani-Pichavant^24^, Karen Salomon-Escoto^14^, Jennifer Seifert^7^, Kamil Slowikowski^9,13^, Darren Tabechian^8^, Jason D. Turner^11^, Paul J. Utz^24^, Gerald F. M. Watts^9^, Kevin Wei^9^

^8^University of Rochester Medical Center, Rochester, NY, USA. ^9^Brigham and Women’s Hospital and Harvard Medical School, Boston, MA, USA. ^10^University of California, San Diego, La Jolla, CA, USA. ^11^University Hospitals Birmingham NHS Foundation Trust and University of Birmingham, Birmingham, UK. ^12^Benaroya Research Institute at Virginia Mason, Seattle, WA, USA. ^13^Broad Institute of MIT and Harvard, Cambridge, MA, USA. ^14^University of Massachusetts Medical School, Worcester, MA, USA. ^15^Feinstein Institute for Medical Research, Northwell Health, Manhasset, New York, NY, USA. ^16^Oklahoma Medical Research Foundation, Oklahoma City, OK, USA. ^17^University of Alabama at Birmingham, Birmingham, AL, USA. ^18^Weill Cornell Graduate School of Medical Sciences, New York, NY, USA. ^19^Barts Health NHS Trust, London, UK. ^20^Northwestern University Feinberg School of Medicine, Chicago, IL, USA. ^21^University of Pittsburgh School of Medicine, Pittsburgh, PA, USA. ^22^Graduate School of Medical and Dental Sciences, Tokyo Medical and Dental University, Tokyo, Japan. ^23^Arthritis Research UK Centre for Genetics and Genomics, Centre for Musculoskeletal Research, The University of Manchester, Manchester, UK. ^24^Stanford University School of Medicine, Palo Alto, CA, USA.

## Author contributions

Designing research studies: ZB, NB, MA, AM, REM, SG, FW, DEO Designing and implementing algorithms and frameworks: ZB

Acquiring data: MA, CH, EAM, NEB, SP, ES, ED, MHS, MJL, SS, CP, AM, REM, SG, FW, DEO

Conducting experiments: ZB, MA, EAM, NEB, SP, AM, REM, FW, DEO

Analyzing data: ZB, NB, MA, CH, EAM, NEB, SP, ES, ED, MHS, MOF, CSJ, HZ, MJL, SS, CP, AM, REM, FZ, SG, FW, DEO

Writing the manuscript: ZB, NB, MA, CH, EAM, NEB, SP, ES, ED, MHS, MOF, CSJ, HZ,MJL, SS, CP, AM, REM, FZ, SG, FW, DEO

## Competing interests

Authors declare that they have no competing interests.

## Data and materials availability

All data sources are available in the main text or the supplementary materials. For codes and any questions, please contact Zilong Bai at.

## Supplementary Materials

### Materials and Methods

#### Identify differentially expressed genes via ANOVA test on pain-level-based groupings

We investigated the association between gene expression and the grouping structure of patient-reported pain in an attempt to identify individual genes that are significantly associated with pain. The 22 patients from HSS with relatively low inflammation were binned into low, medium, and high pain groups. We conducted the one-way ANOVA test between gene expression and pain-level-based groupings of data. To fully utilize the patient-reported pain data, we determine the thresholds for splitting these 22 patients by different binning strategies applied to the 165 total patients from the HSS dataset with HOOS/KOOS pain scores on record. We investigated three binning strategies: (i) quartile-based: high pain was associated with pain score below the first (lower) quartile, medium pain is defined between the first (lower) and third (upper) quartiles, and low pain with the pain score above the third (upper) quartile. This resulted in 6 patients with high pain, 15 patients with medium pain, and 1 patient with low pain. (ii) 33-percentile-based: the pain scores are grouped into thirds by percentiles, i.e., 0-33, 34-66, 67-100. This resulted in 9 patients with high pain, 12 patients with medium pain, and 1 patient with low pain. (iii) mean/stdev-based: the binning thresholds of pain scores are defined by the mean and standard deviation computed using the pain scores of all 165 patients. Pain scores below the mean - standard deviation encompassed patients with high pain, between mean - standard deviation and mean + standard deviation are the patients with medium pain, and above the mean + standard deviation are the patients with low pain. This resulted in 5 patients with high pain, 17 patients with medium pain, and 0 patients with low pain. We performed one-way ANOVA tests using the aov() package in the R stats library. We applied FDR for multiple testing corrections using the p.adjust() function with default settings to all tests for each ANOVA. No gene demonstrated statistical significance in any of these ANOVA tests after FDR-adjustment (i.e., no gene showed FDR-adjusted p-values < 0.05). Filtered by raw p-value < 0.05, we identified one gene (symbol: FAM178B) associated with pain in the quartile-based grouping, 11 pain-associated genes using the 33-percentile-based grouping (Table S4), and zero pain-associated genes from the mean/stdev-based grouping of patients. We performed Pathway Enrichment Analysis on the differentially expressed genes (i.e., FDR-adjusted p-value <0.05) using g:Profiler (https://biit.cs.ut.ee/gprofiler/) with the default background setting for all ANOVA results with data sources for gene ontology and biological pathways specified. In our percentile-based pain bins, we identified 4 significantly enriched (i.e. adjusted p-values <0.05) pathways: methanethiol oxidase activity, ferritin receptor activity, Sulfur metabolism, and TGF-beta receptor signaling in skeletal dysplasias. The PEA results for the other ANOVA tests yielded no significantly enriched pathways (one differentially expressed gene in quartile-based grouping and zero significant genes in mean/stdev-based pain groupings).

#### Ablation study of GbGMI using alternative approaches for gene prioritization

To demonstrate the impact of using graph structure over patients for gene prioritization, we examined different alternatives to our Laplacian-Score-based approach for gene prioritization in our GbGMI framework to identify pain-associated genes. In particular, we sorted the genes based on the correlation tests - i.e., Pearsons, Kendall’s, and Spearman’s, between individual gene’s expression and pain score, and the one-way ANOVA test between gene expression and the pain-level-based groupings of data - i.e., the quartile-based, 33-percentile-based, mean/stdev-based patient groupings as described in the previous section. The rows (corresponding with genes) in the gene expression matrix are reordered according to one of the alternative gene prioritization approaches. For fair comparison with our main results, the top 815 genes in each of these alternative gene prioritization lists are selected as the pain-associated gene module. The multi-gene expression of each alternative pain-associated gene module is embedded into univariate summary scores for the patients via t-SNE as in our GbGMI. We measure the association between the different summary scores and the pain score by different correlation tests (Table S5). Note that none of these alternative gene prioritization approaches enabled GbGMI to identify statistically significant correlation between its top 815 pain-associated gene module summary score and the pain score (i.e., their p-values ≫ 0.05). On the contrary, our GbGMI using Laplacian-Score for gene prioritization achieved P = 0.0013 ≪ 0.05 in Kendall correlation test between its top-815 pain associated gene module summary score and the pain score.

#### Sensitivity analysis of GbGMI on the HSS low-inflammatory patients

We conducted sensitivity analysis to investigate how the patient composition affects the gene prioritization and pain-associated gene subset identification results on the HSS dataset. Specifically, we subsampled the n=22 low-inflammatory patients with the leave-one-out strategy, resulting in 22 different subsampled datasets. On each subsampled dataset, we applied our GbGMI framework to build its pain-score similarity matrix, compute the Laplacian scores for the m=2,227 low-inflammatory genes for prioritization, and determine the cut-off using t-SNE embeddings and correlation test between summary score and pain score to identify the pain-associated gene subset. The resultant 23 (22 for subsampled and 1 for overall) pain-associated gene prioritization gene lists and pain-associated gene subsets are compared to demonstrate how sensitive our GbGMI is with respect to changes in patient set. Each pair of gene prioritization gene lists are compared via Spearman’s correlation test, with the minimum correlation coefficient ⍴_*min*_ = 0.7472 and all p-values ≪ 0.05. (Fig. S6A for pairwise correlation coefficient heatmap.) Each pair of pain-associated gene subsets are compared using the Fisher’s exact test, with the m=2,227 low-inflammatory genes as the background genes. Note that p-value ≪ 0.05 between the pain-associated gene identification results of each leave-one-out patient subset and the overall relatively low-inflammatory patient set. Only 9 pairs of patient subsets with single patient excluded did not show significant overlap (p-value >0.05), e.g., exclude-RA57.SYN vs exclude-RA147.SYN. (Fig. S6B for pairwise Fisher’s exact test p-value heatmap.)

#### Methodological Validation of GbGMI on the early RA dataset

In this section, we validate the consistency of low-inflammatory pain-associated genes identified across the established RA (i.e., HSS (*9*) and the early RA datasets (i.e., PEAC (*10*)). For fair comparison, the gene list from RNA-seq on PEAC is intersected with the m=2,227 low-inflammatory genes that we identified from the HSS dataset, resulting in a 2,018-gene list for low-inflammatory genes on PEAC. Among the 2,018 PEAC low-inflammatory genes, 738 were identified to be pain-associated by GbGMI from the HSS dataset. To directly identify pain-associated genes with GbGMI from the 2,018 PEAC low-inflammatory genes, we grouped the samples labeled as fibroid or ungraded early RA subset and formed a low-inflammatory subset of n=22 patients. GbGMI was applied to this 2,018 × 22 PEAC low-inflammatory gene expression matrix and identified a 658-gene module significantly associated with pain measured by VAS score. The 738 HSS and the 658 PEAC pain associated genes demonstrated significant overlap (p-value = 0.0016 and 95% CI [1.1215, 1.6598]) in Fisher’s exact test with the 2,018 PEAC low-inflammatory genes as background).

#### Ligand-receptor analysis using mouse nervous system scRNA-seq

We also performed ligand-receptor interaction analysis between the pain-associated genes expressed by the four synovial fibroblast subtypes (CD34+ sublining fibroblasts [SC-F1], HLA-DRAhi sublining fibroblasts [SC-F2], DKK3+ sublining fibroblasts [SC-F3], and CD55+ lining fibroblasts [SC-F4]) (*18*) and a mouse nervous system scRNAseq dataset from an adolescent mouse nervous system dataset (*73*). We predicted considerable connections between lining fibroblasts (SC-F4) and peripheral sensory neurofilament, peptidergic and non-peptidergic neurons, as well as sympathetic cholinergic and noradrenergic neurons (Fig. S7). As we will detail in the following, we considered three different scenarios to identify differentially expressed genes in a specific neuronal subpopulation from the mouse nervous system scRNA-seq data: each neuronal cell type vs all the other cell types (including non-neurons), each neuronal cell type vs the other neurons, and comparison among three different specific neuron subtypes (i.e., peripheral sensory neurofilament neurons, peripheral sensory peptidergic neurons, and peripheral sensory non-peptidergic neurons) (Fig. S7).

##### Data

We used the scRNA-seq data from a mouse nervous system dataset (*73*) to generate the transcriptome profile of individual cell-types in the adolescent mouse nervous system. Our analysis used the expression values and metadata for each subpopulation of component cells provided by the original publication. The raw sequence data is deposited in the sequence read archive under accession SRP135960 in the National Center for Biotechnology Information (NCBI) library. The cell subpopulations identified in this mouse nervous system scRNA-seq dataset comprises subpopulations of cells (mainly neurons), from both central (CNS) and peripheral nervous systems (PNS) (*73*).

##### Method

We predicted interactomes for ordered pairs of tissues or cell types that we investigate based on the overall interactome containing more than 3,000 interactions (denoted *I*^*o*^) built by (*19*). We identified ligand-to-receptor unidirectional signaling from one tissue or cell type to another. To identify the interactome for a specific ordered pair of tissue or cell types, the ligand-encoding genes from *I*^*o*^were intersected with the genes that satisfy the ligand-side inclusion criteria in all samples/cells in a specific tissue or cell type wherein we investigate the upstream component of the signaling, and similarly the receptor-encoding genes of *I*^*o*^were intersected with the genes satisfying receptor-side inclusion criteria in another specific tissue or cell type wherein we investigate the downstream component of the signaling.

We generated the interactomes between each of the 4 synovial fibroblast subtypes (*18*) and each selected specific neuronal subpopulation of the mouse nervous system (*73*). On the mouse nervous system scRNA-seq data, we considered three different scenarios to identify differentially expressed genes in a specific neuronal subpopulation: (a) each neuronal cell type vs all the other cell types (including non-neurons), (b) each neuronal cell type vs the other neurons, and (c) comparison among three different specific neuron subtypes (i.e., peripheral sensory neurofilament neurons, peripheral sensory peptidergic neurons, and peripheral sensory non-peptidergic neurons). In each scenario, the differentially expressed genes (i.e., adjusted p-value <0.05) with higher expression values in a specific neuronal cell type are identified by the FindMarkers function from the R package Seurat (*72*) to be the marker genes of a specific neuronal subpopulation. We present the two unidirectional interactomes of each scenario in Fig. S7, where we also summarize the quantity of ligand-receptor interactions associated with each specific fibroblast or neuronal cell type, as well as the total number of interactions, in each tissue-wise direction.

## Supplementary Figures

**Fig. S1.**
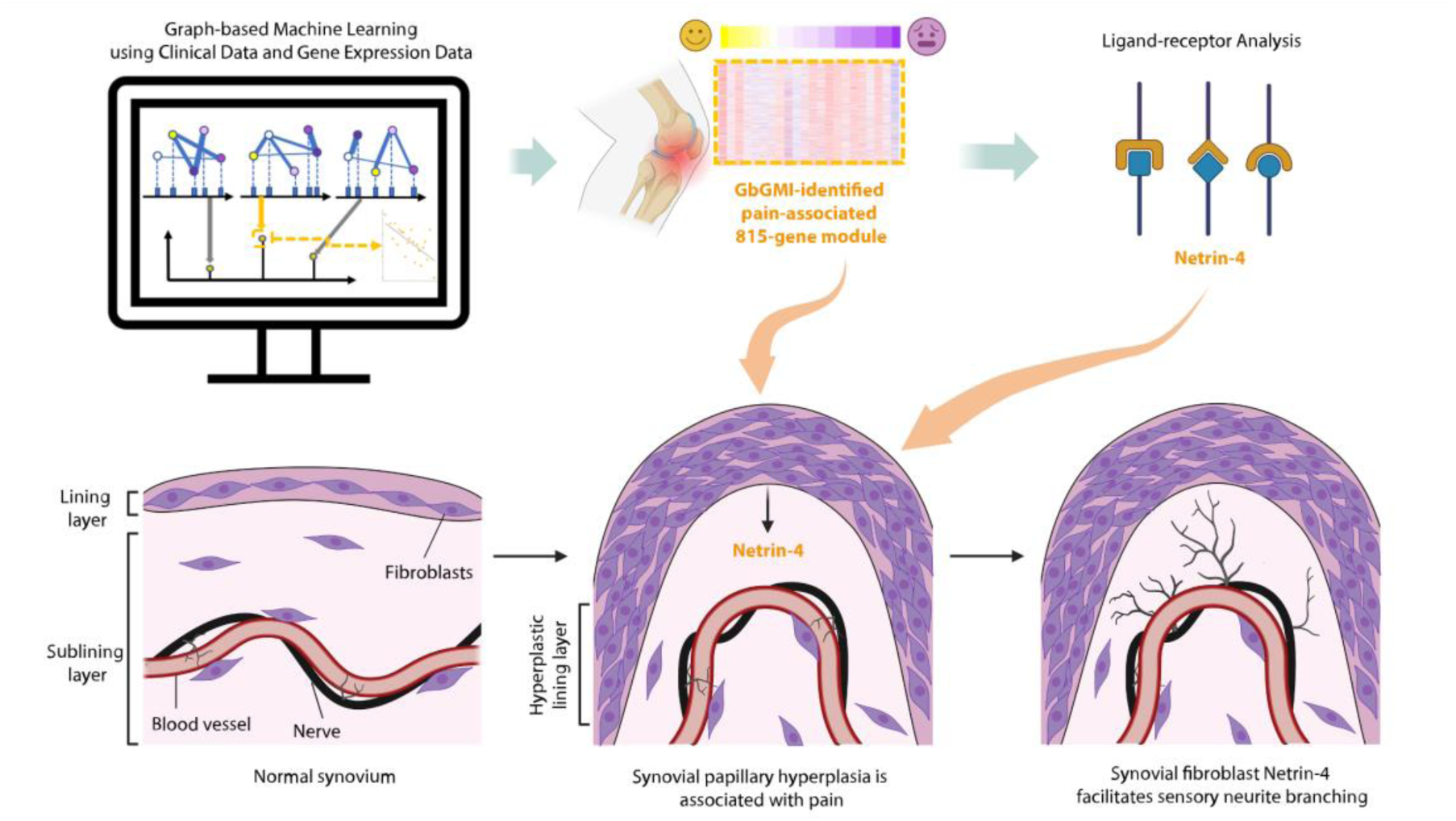
Overview of Study. Graph-based Machine Learning with Clinical Data and Gene Expression Data Identified Synovial Fibroblast Genes Associated with Pain which Affect Sensory Nerve Growth in Rheumatoid Arthritis.

**Fig. S2.**
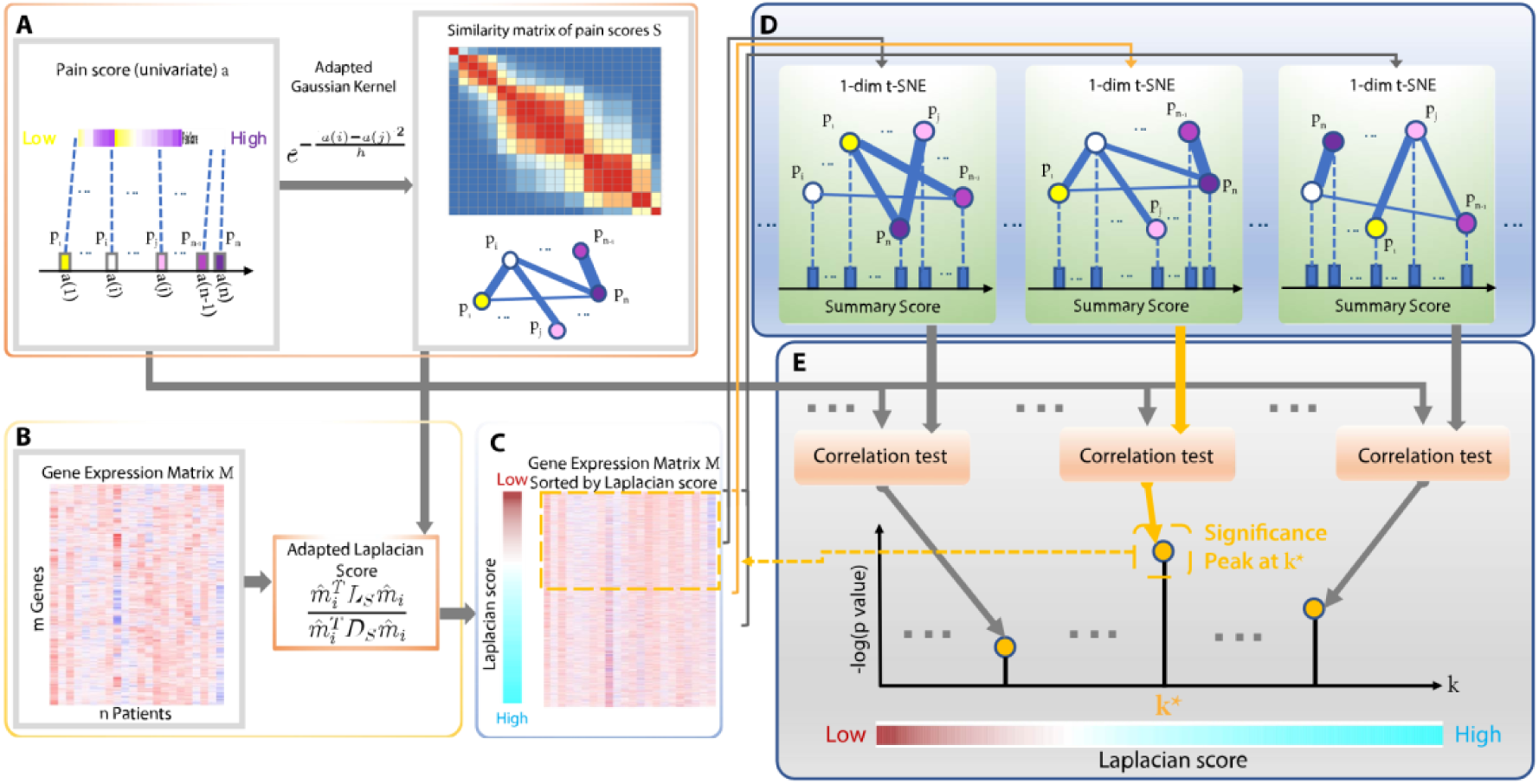
A diagram illustrating the framework of GbGMI for identifying a synovial gene expression signature that associates with pain. (**A**) Construction of similarity matrix over patients based on a univariate patient-level attribute (e.g., HOOS/KOOS pain score). (**B**) Computing Laplacian score for each gene by measuring the concordance between its expression values and the similarity matrix *S* of pain scores from the same set of patients. (**C**) Synovial gene expression matrix with genes sorted by their Laplacian scores in ascending order to form the gene prioritization list ℒ_*s*_, according to which the rows of the gene expression matrix *M* are reordered. (**D**) Generating the gene expression summary score vector *s*_*k*_ for each candidate group of top k genes from ℒ_*s*_ by performing the 1-dim t-SNE embedding on their *k* × *n* gene expression submatrix *M*_1:*k*,∗_. (**E**) Performing correlation tests between the gene expression summary score *s*_*k*_ and the input univariate pain score *a* over the same patients. The *k*^∗^where the significance of correlation coefficients peaks is used to select the group of top *k*^∗^genes from the prioritization list ℒ_*s*_ as the synovial gene expression signature that is significantly associated with pain.

**Fig. S3.**
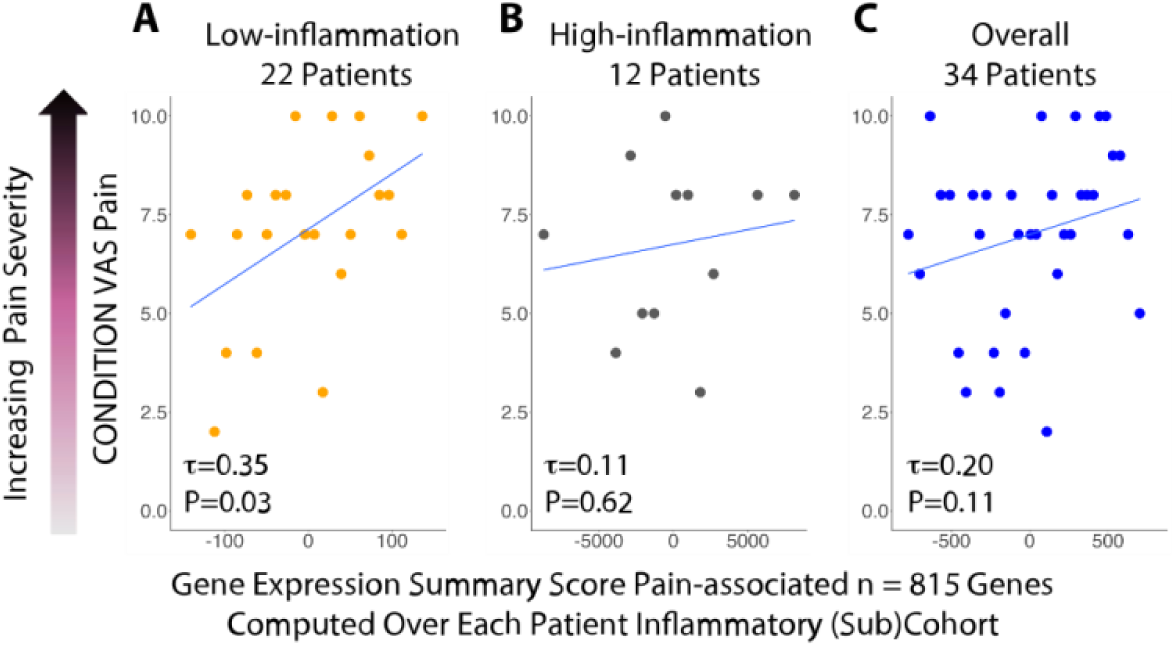
GbGMI-identified pain-associated synovial gene expression compared against VAS pain scores in patients with established RA at different levels of synovial inflammation. (**A**), (**B**), and (**C**): Condition-caused VAS pain score according to the summary score of the 815 GbGMI-identified genes in patients with low synovial inflammation (n=22), high synovial inflammation (n=12), and overall patients with different levels of synovial inflammation (n=34) respectively.

**Fig. S4.**
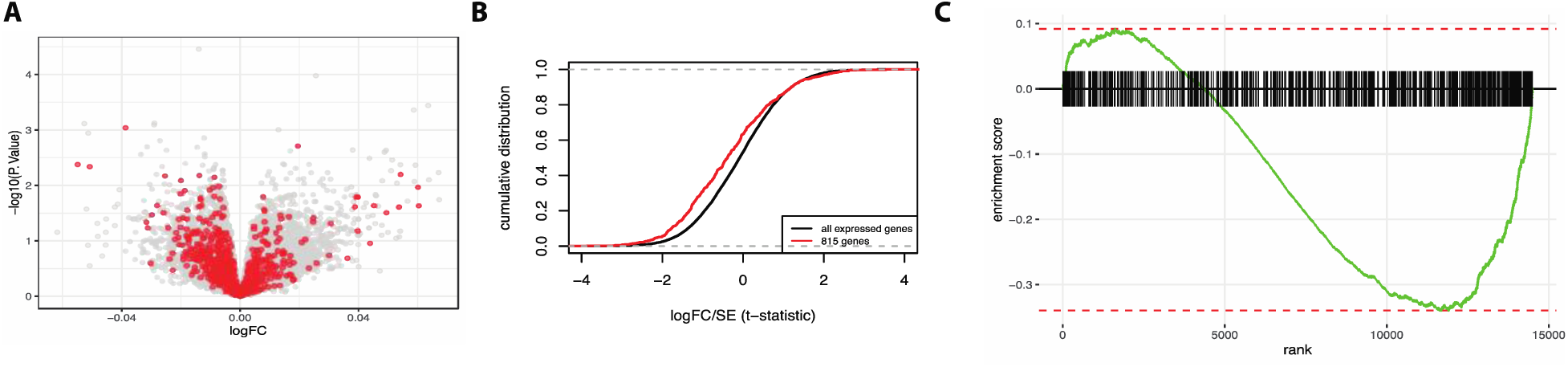
GbGMI-identified pain-associated gene expression increases with increasing pain severity. (**A**) Volcano plot,-log10(nominal p value) and LogFC(Pain score), of differential expression analysis with LIMMA (*14*) for all genes (grey dots) and 815 GbGMI-identified pain-associated genes (red dots) in the bulk RNA-seq data (*9*). (**B**) Cumulative distribution versus t-statistic comparing the 815-gene set and the set of all genes in synovial tissue gene expression data (*9*). (**C**) GSEA querying the 815 pain-associated genes on the genes from the expression data sorted by differential expression analysis (see Materials and Methods). Analysis of GbGMI-identified genes in (**A**), (**B**), and (**C**) consistently indicated that the pain-associated 815 genes as a group decreased expression as the HOOS/KOOS pain score increases, hence indicating their positive correlation with pain severity.

**Fig. S5.**
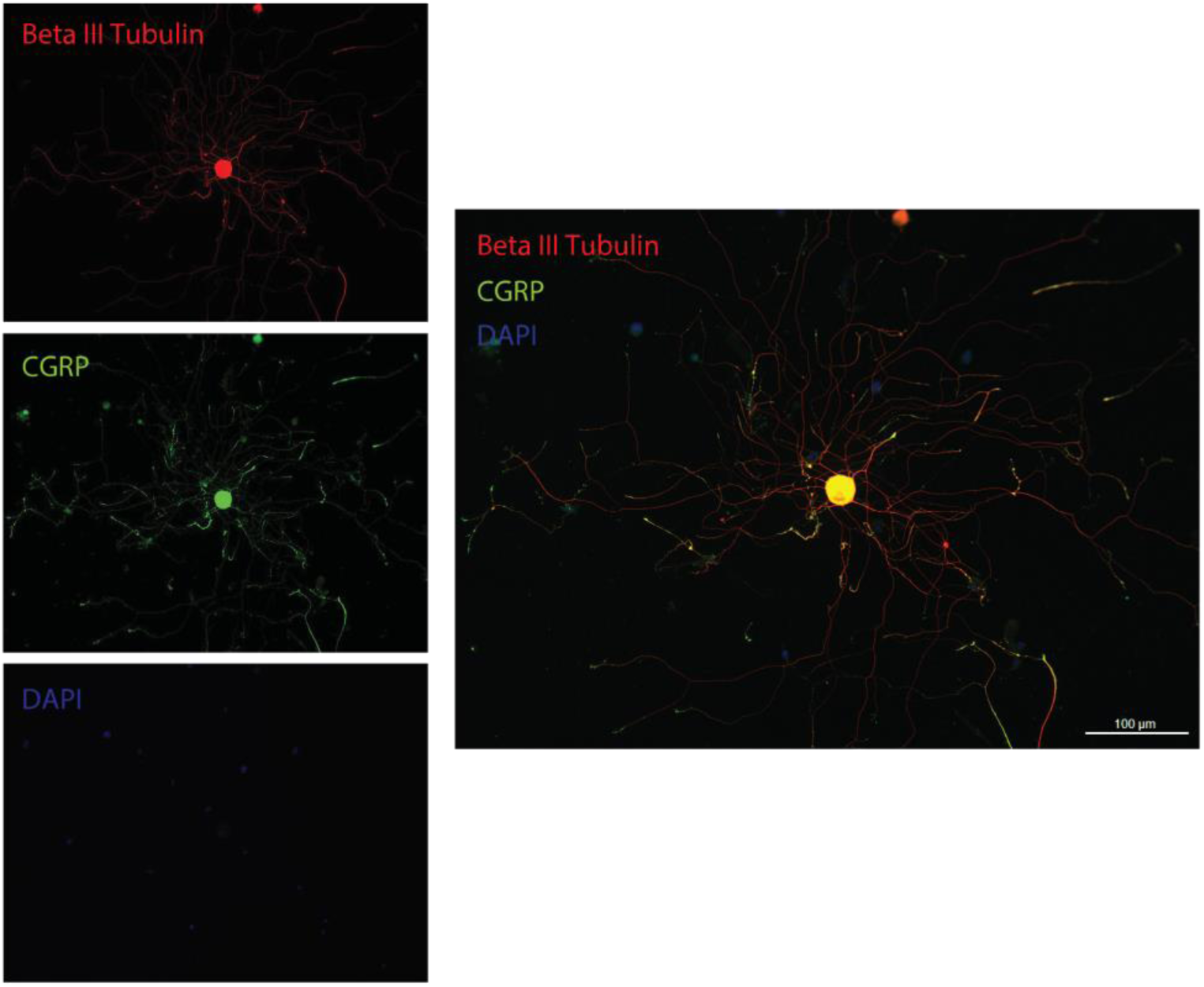
Representative staining of CGRP+ DRG neuron *in vitro*.

**Fig. S6.**
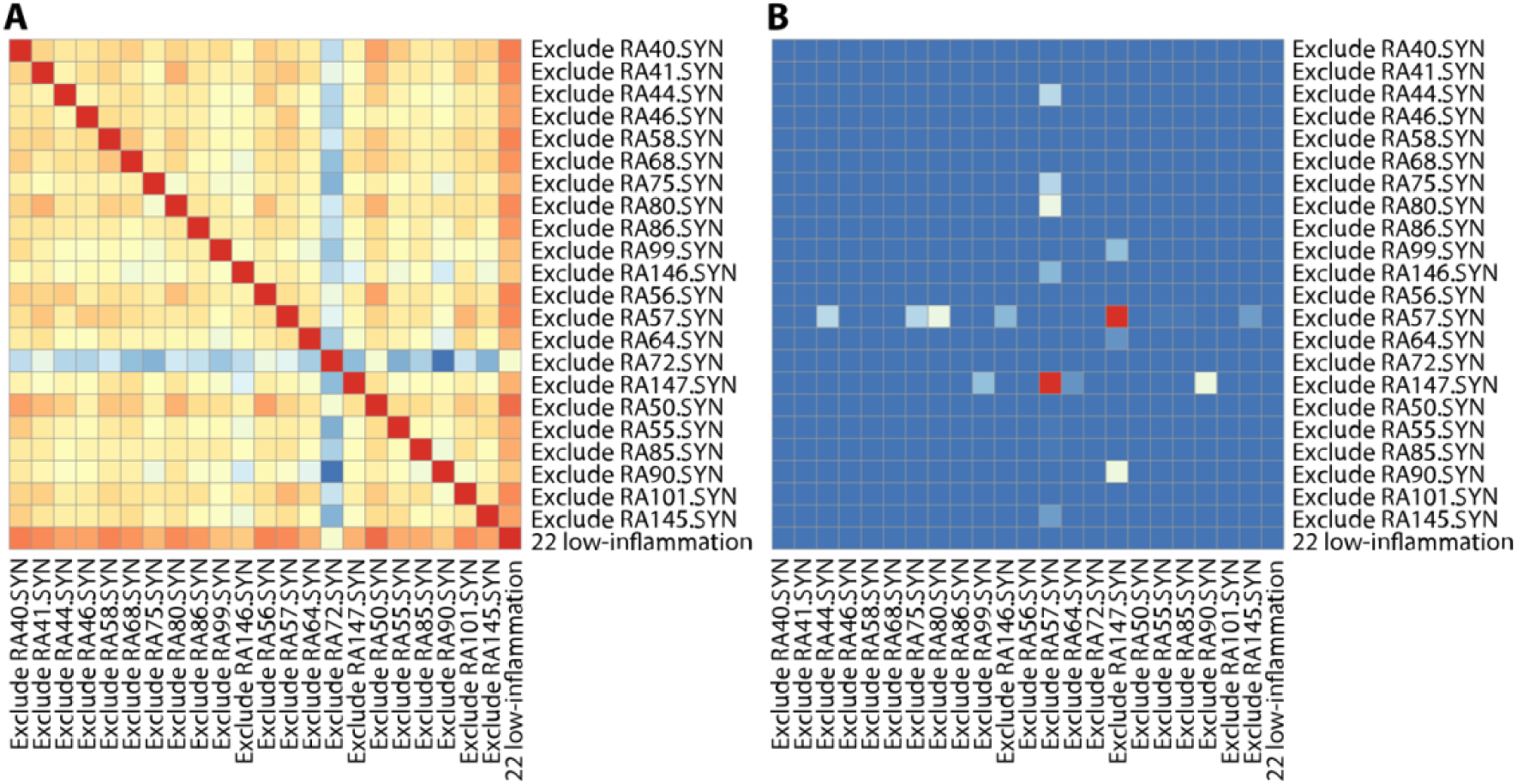
Sensitivity analysis of GbGMI on the HSS low-inflammatory patients. (**A**) Heatmap of Spearman correlation coefficient between each pair of Laplacian-Score-based gene prioritization lists generated using different patient subsets. (**B**) Heatmap of Fisher’s exact test p-values between the pain-associated gene module identification results of different patient subsets. In both (**A**) and (**B**), the first 22 rows/columns indicate the leave-one-out patient subsets, the last rows/columns indicate all the 22 patients with relatively low inflammation from the HSS cohort.

**Fig. S7.**
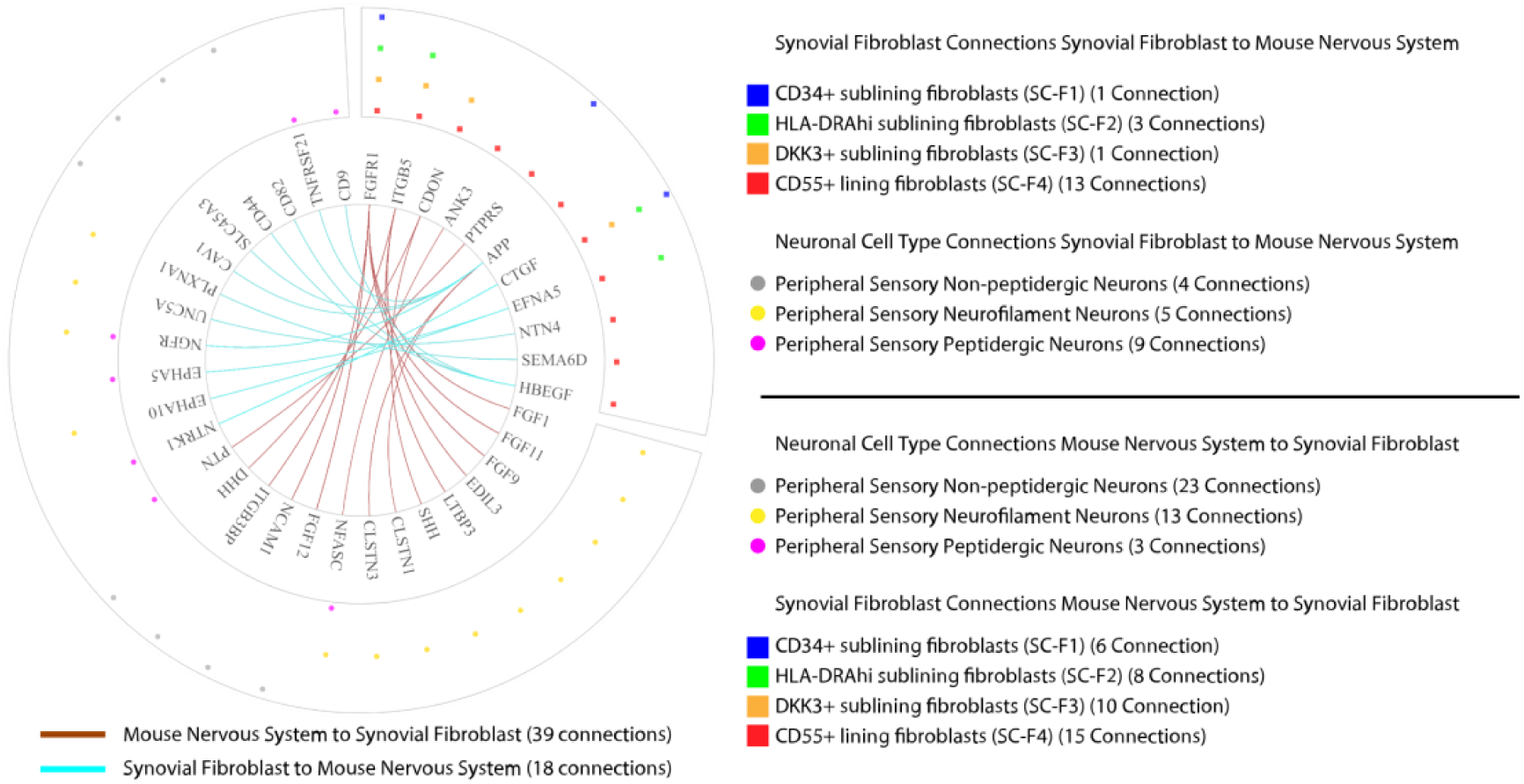
Potential ligand-receptor interactions between synovial fibroblasts and neurons in the mouse nervous system. The circos plot shows the two unidirectional interactomes between the four synovial fibroblast subtypes and a subset of neuronal cell subpopulations from the mouse nervous system. The marker genes of its neuron subtypes are identified through comparison among three different specific neuron subtypes (i.e., peripheral sensory neurofilament neurons, peripheral sensory peptidergic neurons, and peripheral sensory non-peptidergic neurons). The outermost circles indicate the RA synovial fibroblast subtype of the cells or mouse nervous system neuronal cell subpopulation expressing corresponding ligand or receptor genes. The middle layer shows whether a gene is ligand-coding or receptor-coding in its associated interactions. The inner layer contains gene names. The two tissue-wise directions are distinguished by the colors of connections between gene names. The number of connections associated with the ligand/receptor genes in each fibroblast subtype or neuron subtype and in each unidirectional tissue-wise relation are summarized in the corresponding legends.

**Table S1:** Pathway Analysis (Data file S1) Pathway enrichment analysis for the GbGMI identified 815-gene pain-associated genes. Modified Fisher’s exact test based on the hypergeometric distribution; -log10 of P-values after multiple testing correction with g:SCS (*65*) are color-coded for the GO: Biological Processes (BP) terms (adjusted P-value < 0.05)

**Table S2:** Fibroblast–DRG receptor ligand interaction analysis (Data file S2) The ligand-receptor pairs identified between the pain-associated genes expressed by four synovial fibroblast subtypes in human RA synovial tissue and expressed genes in a human DRG bulk RNA-seq dataset (*20*). Ligand-coding and receptor-coding gene symbols are in the “ligand_gname” and “receptor_gname” columns respectively. The RA fibroblast subtypes of the ligand-coding genes are annotated in the column “ligand_cell_type”. The “receptor_tissue” is human DRG.

**Table S3:** Fibroblast-Sensory nerve subset receptor ligand interaction analysis (Data file S3) The ligand-receptor pairs identified between the pain-associated genes expressed by the synovial fibroblast subsets (*18*) and a mouse scRNA-seq DRG dataset (*21*). Ligand-coding and receptor-coding gene symbols are in the “ligand_gname” and “receptor_gname” columns respectively. The receptor coding genes symbols are translated from mouse gene symbols to their human gene symbol counterparts. The “receptor_cell_type” column denotes the neuronal cell types from the mDRG dataset of each receptor-coding gene. The column “ligand_cell_type” denotes the RA fibroblast subtype of each ligand-coding gene.

**Table S4.** Differentially expressed genes (FDR-adjusted p-value<0.05) identified by ANOVA test using the pain-score percentile-based patient grouping.

| Gene | FDR Adjusted Pval |
| --- | --- |
| SELENBP1 | 0.024 |
| FAM178B | 0.024 |
| MAP7D2 | 0.048 |
| SLC22A17 | 0.048 |
| ZNF423 | 0.048 |
| PLEKHA4 | 0.049 |
| KHDRBS3 | 0.048 |
| LTBP3 | 0.049 |
| SCARA5 | 0.048 |
| AC106786.1 | 0.048 |
| CTD-3049M7.1 | 0.048 |

**Table S5.** Correlation tests between the top 815-gene modules identified by GbGMI using different alternative gene prioritization approaches as alternatives to our Laplacian-Score approach and the pain score. The p-values of Kendall’s, Pearson’s, and Spearman’s correlation tests between the top 815-gene module summary scores yielded by the different gene prioritization approaches and the pain score are presented.

| Alternative gene prioritization approach | P-value of correlation test between top 815-gene module summary score and pain score |  |  |
| --- | --- | --- | --- |
|  | Kendall | Pearson | Spearman |
| Kendall's correlation | 0.6509 | 0.5643 | 0.6905 |
| Pearson's correlation | 0.6107 | 0.8613 | 0.7053 |
| Spearman's correlation | 0.9099 | 0.446 | 0.8083 |
| ANOVA test (quantile-based patient grouping) | 0.7343 | 0.9247 | 0.6979 |
| ANOVA test (33-percentile-based patient grouping) | 0.4284 | 0.5960 | 0.3686 |
| ANOVA test (mean/stdev-based patient grouping) | 0.7343 | 0.7613 | 0.8375 |

## Notes

### Competing Interest Statement

The authors have declared no competing interest.

### Funding Statement

National Science Foundation 1750326
National Institute of Arthritis and Musculoskeletal and Skin Diseases NIH R01 AR078268
National Institute of Arthritis and Musculoskeletal and Skin Diseases UC2AR081025
National Institute of Arthritis and Musculoskeletal and Skin Diseases R01AR077019

### Author Declarations

This study was approved by the HSS Institutional Review Board (approval no. 2014-233), the Rockefeller University Institutional Review Board (approval no. DOR0822), and the Biomedical Research Alliance of New York (approval no. 15-08-114-385).

